# Multiple Organ Dysfunction Trajectory Phenotypes After Severe Trauma Are Associated With Distinct Immune Profiles and Outcomes

**DOI:** 10.64898/2026.10.15.26365695

**Authors:** Haiyuan Ding, Madison Cox, Qian Qiu, Nina Mirabadi, David Bobrovnikov, Krista Gerbino, Kelsey Scherler, Kathie-Anne Walters, Grant O’Keefe, Scott C. Brakenridge

**Author notes:** **Corresponding Author: Scott C. Brakenridge, MD**, Harborview Medical Center, Box 359796, 325 Ninth Ave, Seattle, WA 98104. Supported in part by the National Institutes of Health, National Institute of General Medical Sciences (NIGMS) grants R35GM142481 and T32GM121290. All authors report no conflict of interest.

## Abstract

**BACKGROUND:** Traditional organ dysfunction classifications after severe trauma rely on binary categories (e.g., chronic critical illness [CCI] versus rapid recovery), which likely oversimplify the true heterogeneity of postinjury organ dysfunction trajectories and their underlying immune-biologic drivers. We hypothesized that severely injured trauma patients demonstrate multiple, distinct organ dysfunction phenotypes distinguished by unique immune signatures.

**METHODS:** We performed a prospective, longitudinal, observational cohort study of severely injured trauma patients at high risk for multiple organ dysfunction at a single Level 1 trauma center. Organ dysfunction was quantified daily through 28 days, death, or discharge using a trauma-specific modified Sequential Organ Failure Assessment (mSOFA) score excluding the neurologic subdomain to isolate systemic organ dysfunction from traumatic brain injury. We utilized group-based trajectory modeling to identify longitudinal mSOFA phenotypes. Plasma immune protein trajectories were compared across phenotypes using mixed-effects models with time interaction.

**RESULTS:** Sixty-nine patients were enrolled. Trajectory modeling identified five mSOFA phenotypes (c1-c5) with differences in resuscitation requirements, shock severity, and 28-day mortality, but not baseline demographics, injury severity, or early physiologic parameters. The phenotypes revealed organ dysfunction heterogeneity not fully captured by CCI/rapid recovery classification. Forty of 92 measured immune markers showed phenotype-differentiated trajectories (FDR<0.1). The most fulminant phenotype (c5) was distinguished by IL-17C, CXCL5, and CXCL6, implicating epithelial-barrier and neutrophil-trafficking mechanisms. The phenotype with highest CCI incidence (c3) was distinguished by rising CXCL9, ARTN, and TWEAK, implicating Th1 polarization and tissue-inflammation/repair signaling. The clinically distinct, older-age phenotype (c4) carried the largest differentiating signature (29 proteins), spanning renal injury/repair, endothelial activation, and adaptive lymphocyte modulation.

**CONCLUSIONS:** Organ dysfunction after severe trauma is more biologically heterogeneous than traditional clinical classifications suggest. Distinct mSOFA trajectory phenotypes correspond to distinct immune signatures, supporting separable pathobiology rather than a single axis of injury severity, with implications for phenotype-targeted precision immunomodulatory approaches in critical illness.

**LEVEL OF EVIDENCE:** Level III, Prognostic/Epidemiological.

## BACKGROUND

Advances in resuscitation and critical care have progressively reduced early mortality after severe trauma. Among survivors, the trajectory of organ dysfunction, not merely its presence, is a dominant determinant of outcome, and a growing proportion of patients survive to chronic critical illness (CCI), marked by persistent organ dysfunction and dismal long-term outcomes.(1–3) This clinical heterogeneity is thought to reflect mechanistic heterogeneity in the underlying host immune response. Severe injury provokes a rapid, simultaneous dysregulation of innate and adaptive immunity, a “genomic storm” of variable magnitude, composition, and resolution kinetics between patients.(4–6) We have previously shown that trajectory-based immune biomarker clustering identifies distinct immune endotypes among severely injured patients that track with clinical course and mortality.(7) However, this host response heterogeneity remains incompletely understood, limiting the ability to predict individual clinical trajectories or implement effective precision medicine approaches to organ dysfunction after severe injury.

Recent trauma studies applying trajectory modeling and clustering approaches to the Sequential Organ Failure Assessment (SOFA) score have demonstrated that post-injury organ dysfunction is more heterogeneous than binary complicated/uncomplicated or CCI/rapid-recovery classification suggests.(8, 9) However, standard SOFA scoring embeds a neurologic (Glasgow Coma Scale) subdomain that confounds systemic immune-driven organ dysfunction with primary traumatic brain injury, both studies included large numbers of patients not at high risk (i.e., minimal/absent shock) of persistent organ dysfunction related outcomes, and neither study paired the derived phenotypes with concurrent immune biology. A recent multi-omic study by Cohen *et al* inverted this approach, deriving molecular endotypes from longitudinal plasma proteomic/metabolomic trajectories with organ dysfunction outcomes as a downstream correlate. (10) However, there still remains a significant gap of knowledge in understanding the breadth of heterogeneity of organ dysfunction trajectories and their driving immunologic mechanisms after severe trauma with hemorrhagic shock.

We hypothesized that among a cohort of severely injured trauma patients with hemorrhagic shock and similar clinical characteristics: 1) there are multiple, unique phenotypes of longitudinal organ dysfunction trajectory, and 2) these phenotypes exhibit distinct immune biologic profiles suggesting differences in underlying biologic mechanisms.

## METHODS

### Study design, inclusion/exclusion criteria and enrollment

This analysis is part of a prospective, longitudinal, observational cohort study of severe trauma patients at a single Level 1 trauma center. The parent study enrolled critically ill trauma patients at high risk for multiple organ dysfunction and CCI, with longitudinal multi-omic immune profiling, clinical data collection, and outcomes assessment, with the primary aim of characterizing post-injury organ dysfunction heterogeneity by identifying distinct trajectories (phenotypes) and their underlying biologic mechanisms (endotypes). Inclusion criteria included patients admitted to the Trauma ICU service with an injury severity score (ISS) ≥25 and/or evidence of hemorrhagic shock (systolic blood pressure <90 mmHg or lactate >2 mmol/L), adapted from the Inflammation and Host Response to Injury program and our prior cohorts.(2, 7) Potential enrollees were prospectively adjudicated by the principal investigator (**SDC 2 Expanded Methods**). Exclusion criteria included the following: 1) patients not expected to survive >48 hours, 2) investigator-adjudicated, neurologically devastating traumatic brain injury, 3) prisoners or pregnant patients, 4) patients receiving chronic corticosteroids, immunosuppressive therapies, or previous bone marrow transplantation, 5) or severe/confounding medical comorbidities including pre-existing end-stage renal disease, severe congestive heart failure (NY Heart Association Class IV), chronic liver disease with MELD score ≥15, HIV infection with CD4+ count <200 cells/mm3, or active/symptomatic COVID-19 virus infection. Informed consent was obtained from the patient, or proxy if incapacitated (see **SDC2 Expanded Methods**). This study was approved in full compliance with the Declaration of Helsinki and considered as “minimal risk” human subjects research by the University of *Washington* Institutional Review Board (IRB).

### Biospecimen collection and immune proteomic analyses

Peripheral whole blood was collected from enrolled trauma patients within 48 hours of admission, and at days 4, 7, 10, 14, 21 and 28 or until death/hospital discharge. Blood was collected into Na-Heparin tubes and processed within 1 hour of collection. Plasma was isolated by centrifugation at 1800*g* for 10 min, aliquoted, and stored at –80°C for batched proteomic analysis. Select immune-relevant plasma proteins were quantified using the Olink® Target 96 Inflammation panel (Olink Proteomics, Uppsala, Sweden) following the manufacturer’s protocol. Cryopreserved plasma samples were thawed on ice, clarified by centrifugation, and assayed in duplicate. Data processing was performed with Olink NPX Manager, including background correction and normalization to extension and inter-plate controls. Protein abundance is reported as Normalized Protein eXpression (NPX, log2 scale) values. Protein values below the limit of detection in >25% of samples were excluded prior to analysis.

### Organ dysfunction assessment and quantification

Post-injury organ dysfunction was quantified using a trauma-relevant, modified SOFA (mSOFA) score schema (**SDC2 Expanded Methods**). This mSOFA score calculation is based on standard SOFA score criteria(11) with exclusion of the Glasgow Coma Scale (GCS)/central nervous system (CNS) organ system score. Eliminating the CNS component of the SOFA score removes the confounding effect of traumatic brain injury (TBI) on post-injury multi-organ dysfunction quantification, as previously described.(4, 12, 13) Additionally, the mSOFA score incorporates organ support criteria that reflect current critical care support modalities and practices (**SDC2 Expanded Methods**).(14) All components of the mSOFA score are calculated from ICU clinical flowsheet and laboratory data contained within the institutional electronic medical record (EMR). For each subject, daily mSOFA individual organ system and total scores were calculated to quantify and trend organ dysfunction over time. Conservative imputation strategies were delineated *a priori* for the most common missing organ subscore variables (Hepatic, Respiratory; **SDC2 Expanded Methods**).

### Clinical data collection and definitions of clinical outcomes

Baseline clinical characteristics, including age, sex, comorbidities, mechanism and severity of injury, operative procedures, blood product transfusion, and shock severity, were collected via a combination of prospective clinical chart review and batched EMR data collection. Patients were prospectively tracked until hospital discharge or death and were categorized by previously described “coarse” organ recovery clinical outcomes at day 14 as rapid recovery (RR) or chronic critical illness (CCI). (2) Rapid recovery (RR) patients were defined as those downgraded from ICU level care within 14 days of injury with resolution of organ dysfunction. Chronic critically ill patients (CCI) remained in the ICU with persistent organ dysfunction greater than or equal to 14 days. For analysis, patients who died before day 14 (n=3) were grouped with CCI as an adverse coarse outcome.

### Group-based trajectory modeling, phenotype development & statistical analyses

We utilized group-based trajectory modeling (GBTM) to identify longitudinal mSOFA cluster phenotypes, performing latent class trajectory modeling on mSOFA scores over the first 28 days of hospitalization. In the case of death, a mSOFA value of 20 was appended to the patient’s time scores, and in the case of discharge, a mSOFA value of –1 was appended.(15) Clustering was performed using the “lcmm” package (v2.2.2) in R (v4.6.0). Akaike Information Criterion (AIC), Bayesian Information Criterion (BIC), and log likelihood metrics, along with consideration of cluster group sizes, aided in the manual selection of a second order polynomial model [SOFA ∼ day + I(day^2)] with five outcome clusters (mSOFA phenotypes, c1-c5), all showing high classification certainty (average posterior probability of assignment [APPA], range 0.93-0.99; **SDC2 Expanded Methods**). Demographic and outcome variables of interest, including age, Injury Severity Score (ISS), lactate levels, and transfusion statistics, were compared across the five mSOFA phenotypes using analysis of variance (aov), Mann-Whitney, c^2^ or Fisher’s exact tests in R, as appropriate. For these comparisons, an unadjusted P value <0.05 was considered statistically significant.

In order to identify differences in immune mediator patterns across phenotypes, plasma immune protein abundance values (Olink NPX) were modeled in R against phenotype classifications (c1-c5) over the first 28 days of hospital admission using the “kimma” package (v1.6.3).(16) Models included fixed effects for mSOFA group and time, the interaction between mSOFA group and time, and the random effect of subject ID (i.e., individual patients). Given the exploratory, hypothesis-generating design and moderate panel size (92 analytes) of the proteomic analysis, we applied Benjamini-Hochberg false discovery rate (FDR) correction and considered an FDR <0.1 statistically significant. This threshold represents a balance between the highly conservative 5% convention and more permissive thresholds (up to 20%) for FDR proposed for initial genomic and proteomic discovery-phase analyses.(17–19) All proteins with differences in trajectory at an FDR<0.1 were annotated for function/mechanism by the investigators to standardize subsequent phenotype biological interpretation (**SDC2 Expanded Methods, SDC 3 Table S1**).

### STROBE statement

This study is reported in accordance with the STROBE (Strengthening the Reporting of Observational Studies in Epidemiology) guideline for cohort studies to ensure proper reporting of methods, results, and discussion (**SDC1**).

## RESULTS

### Prospective cohort demographics

The prospective cohort consisted of 69 patients enrolled over an approximately 3-year period (March 2022 to Dec 2024). Patients qualified for enrollment by ISS ≥25 alone (n=13, 18.8%), shock criteria alone (n=10, 14.5%), or both (n=43, 62.3%); three patients (4.3%) enrolled on the basis of preliminary ISS estimation had a final registry ISS <25 without meeting shock criteria. Demographics and clinical characteristics reveal this was a severely injured cohort with evidence of hemorrhagic shock (**Table 1**). All patients (n=69, 100%) underwent at least one operative procedure, and 60 (87.0%) underwent two or more. By coarse clinical outcome trajectory classification, 46% of patients demonstrated a rapid recovery (RR) and 54% developed chronic critical illness (CCI). Notably, there were no differences in baseline demographics, injury characteristics, shock severity or initial 24-hour resuscitation volumes between the RR and CCI clinical outcome groups (**Table 1**).

**Table 1.** Baseline demographic, injury, and shock resuscitation characteristics by clinical trajectory.

| Characteristic | Overall | Rapid Recovery (RR) | Chronic Critical Illness (CCI) | p |
| --- | --- | --- | --- | --- |
|  | (n = 69) | (n = 32) | (n = 37) |  |
| **A. Demographics** |  |  |  |  |
| Age, y, median (IQR) | 46 (35, 57) | 46 (34, 53) | 46 (35, 58) | 0.67 |
| Sex, n (%) |  |  |  | 0.85 |
| Male | 51 (73.9) | 24 (75.0) | 27 (73.0) |  |
| Female | 18 (26.1) | 8 (25.0) | 10 (27.0) |  |
| Race, n (%) |  |  |  |  |
| White | 50 (72.5) | 22 (68.8) | 28 (75.7) | 0.52 |
| Black or African American | 7 (10.1) | 4 (12.5) | 3 (8.1) | 0.70 |
| American Indian or Alaska Native | 3 (4.3) | 0 (0) | 3 (8.1) | 0.24 |
| Native Hawaiian or Other Pacific Islander | 0 (0) | 0 (0) | 0 (0) | --- |
| Asian | 1 (1.4) | 1 (3.1) | 0 (0) | 0.46 |
| Other | 4 (5.8) | 2 (6.3) | 2 (5.4) | 1.00 |
| Unknown | 4 (5.8) | 3 (9.4) | 1 (2.7) | 0.66 |
| Ethnicity, n (%) |  |  |  | 0.88 |
| Non-Hispanic | 59 (85.5) | 27 (84.4) | 32 (86.5) |  |
| Hispanic | 7 (10.1) | 3 (9.4) | 4 (10.8) |  |
| Unknown | 3 (4.3) | 2 (6.3) | 1 (2.7) |  |
| **B. Injury Characteristics** |  |  |  |  |
| ISS, median (IQR) | 30 (26, 41) | 31 (27, 38) | 30 (26, 41) | 0.62 |
| Mechanism, blunt vs penetrating, n (%) |  |  |  | 1.00 |
| Blunt | 61 (88.4) | 28 (87.5) | 33 (89.2) |  |
| Penetrating | 8 (11.6) | 4 (12.5) | 4 (10.8) |  |
| Mechanism of injury, n (%) |  |  |  | 0.34 |
| Fall | 5 (7.2) | 1 (3.1) | 4 (10.8) |  |
| MVC – occupant | 21 (30.4) | 11 (34.4) | 10 (27.0) |  |
| MVC – motorcyclist | 16 (23.2) | 10 (31.3) | 6 (16.2) |  |
| MPC – cyclist (struck or fall) | 3 (4.3) | 0 (0) | 3 (8.1) |  |
| MPC – pedestrian | 15 (21.7) | 6 (18.8) | 9 (24.3) |  |
| GSW | 7 (10.1) | 4 (12.5) | 3 (8.1) |  |
| Penetrating stab wound | 1 (1.4) | 0 (0) | 1 (2.7) |  |
| Other | 1 (1.4) | 0 (0) | 1 (2.7) |  |
| **C. Shock Severity** |  |  |  |  |
| Lowest ED SBP, mmHg, median (IQR) | 82 (68, 96) | 83 (73, 93) | 79 (64, 98) | 0.36 |
| Highest lactate through 6 h, (mmol/L), median (IQR) | 4 (2, 7) | 4 (2, 6) | 5 (3, 8) | 0.20 |
| **D. Initial 24-Hour Resuscitation Volumes** |  |  |  |  |
|  | (n = 69) | (n = 32) | (n = 37) |  |
| Whole blood, mL, median (IQR) | 1500 (500, 3000) | 1750 (1000, 3000) | 1500 (500, 3500) | 0.59 |
| Red blood cells, mL, median (IQR) | 1050 (700, 2275) | 1300 (837, 2275) | 1050 (700, 2446) | 0.76 |
| Fresh frozen plasma, mL, median (IQR) | 923 (481, 1800) | 916 (428, 1749) | 930 (481, 2169) | 0.87 |
| Crystalloid, mL, median (IQR) | 3000 (1500, 4000) | 2800 (1200, 5000) | 3000 (1650, 4000) | 0.86 |

IQR indicates interquartile range; ISS, Injury Severity Score; ED, emergency department; SBP, systolic blood pressure; MVC, motor vehicle collision; MPC, motor/pedal cyclist; GSW, gunshot wound; RR, rapid recovery (including intermediate recovery); CCI, chronic critical illness. P-values represent testing by Mann Whitney U or $\chi^2$ test as appropriate.

### Organ dysfunction trajectory phenotypes

Our analysis identified 5 longitudinal cluster phenotypes (c1-c5) based on mSOFA score trajectories over time (**Figure 1**). Raw data trajectory plots are shown individually across the 5 phenotypes in **SDC4 Figure S1.** Based on organ dysfunction trajectory and relationship to 28-day mortality, we categorized these as lower severity (c1, c2, c3) and higher severity (c4, c5) phenotypes. The lower severity (c1-c3) phenotypes exhibited survival to hospital discharge after progressively slower resolution of organ dysfunction, with c3 having a significantly longer hospital length of stay than c1 and c2 (**Figure 1; SDC4 Figure S2**). All subjects in the higher severity phenotypes (c4, c5; n=12/12, 100%) either developed CCI or died within 28 days of injury. The c4 phenotype exhibited persistent, non-resolving organ dysfunction with late deterioration. The c5 phenotype exhibited a trajectory of fulminant multiple organ failure, with persistent organ dysfunction leading to CCI or death (**Figure 1; SDC4 Figure S1**).

**Figure 1.**
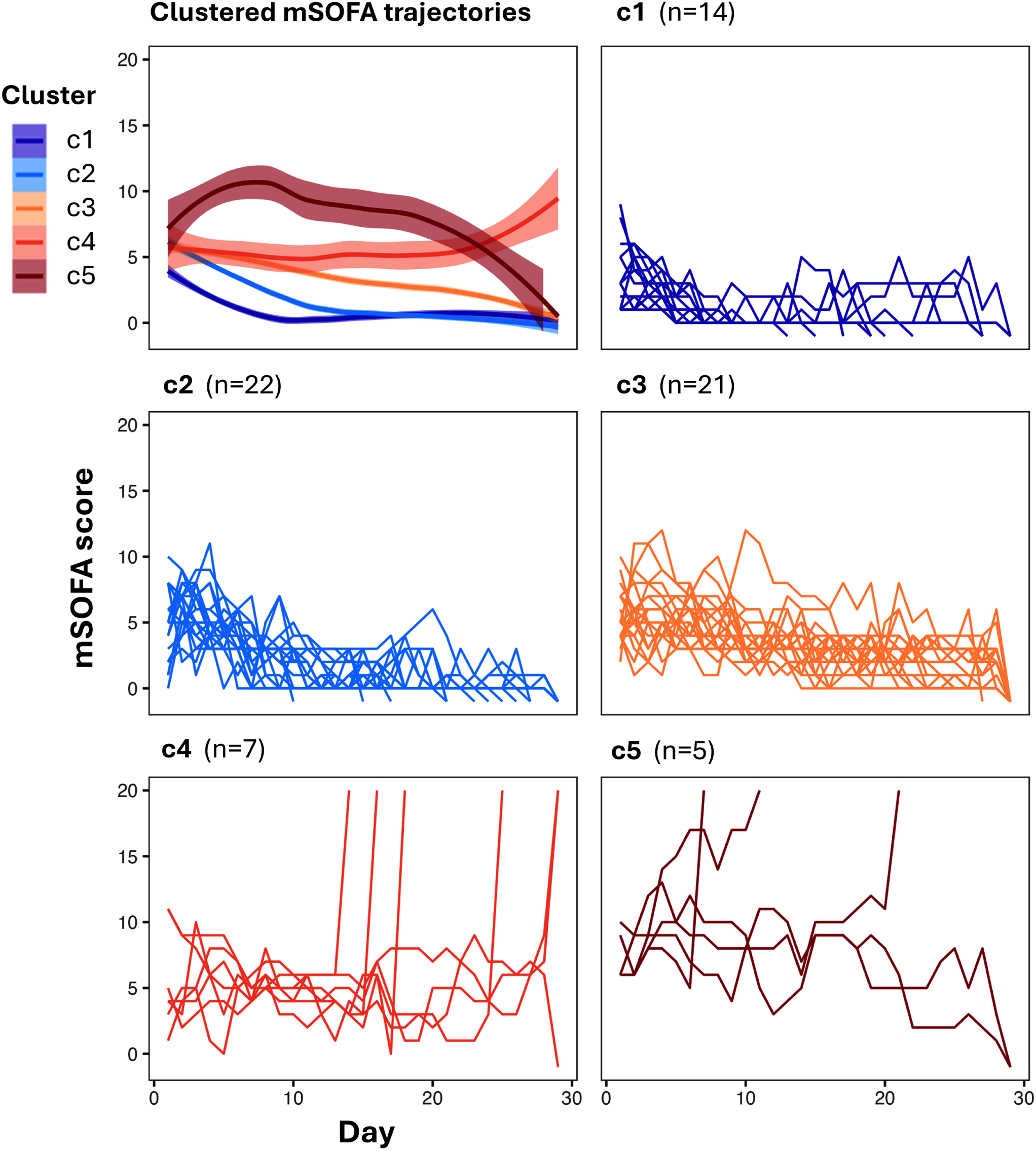
Organ dysfunction trajectory cluster phenotypes after severe traumatic injury. Cluster phenotypes derived from longitudinal mSOFA scoring. Combined smoothed trajectories with standard error are shown out to 28 days (mSOFA trajectories). Cluster sizes: c1 (n=14), c2 (n=22), c3 (n=21), c4 (n=7), c5 (n=5).

Importantly, the mSOFA trajectory phenotypes revealed heterogeneity in organ dysfunction not fully captured by the coarse clinical outcomes (RR/CCI) at day 14, as seen in **Figure 2**. The higher severity phenotypes (c4, c5) represent the subgroup of CCI patients that have the highest risk of 28-day mortality (c4, n=6/7, 86%; and c5, n=3/5, 60%). Three of the 9 deaths occurred before day 14 (c5, n=2; c4, n=1; **SDC4 Figure S1**), and all non-survivors underwent proteomic sampling at each scheduled timepoint until death. However, this heterogeneity extends to the lower severity (c1-c3) phenotypes as well. Although all 57 lower severity phenotype patients (100%) were alive at 28 days, their coarse clinical outcomes at day 14 diverge based upon their organ dysfunction trajectories. While the overwhelming majority of c1 phenotype patients (n=11/14, 79%) exhibit rapid recovery, c2 and c3 phenotypes have mixed, but somewhat skewed coarse clinical outcomes at day 14 (**Figure 2**).

**Figure 2.**
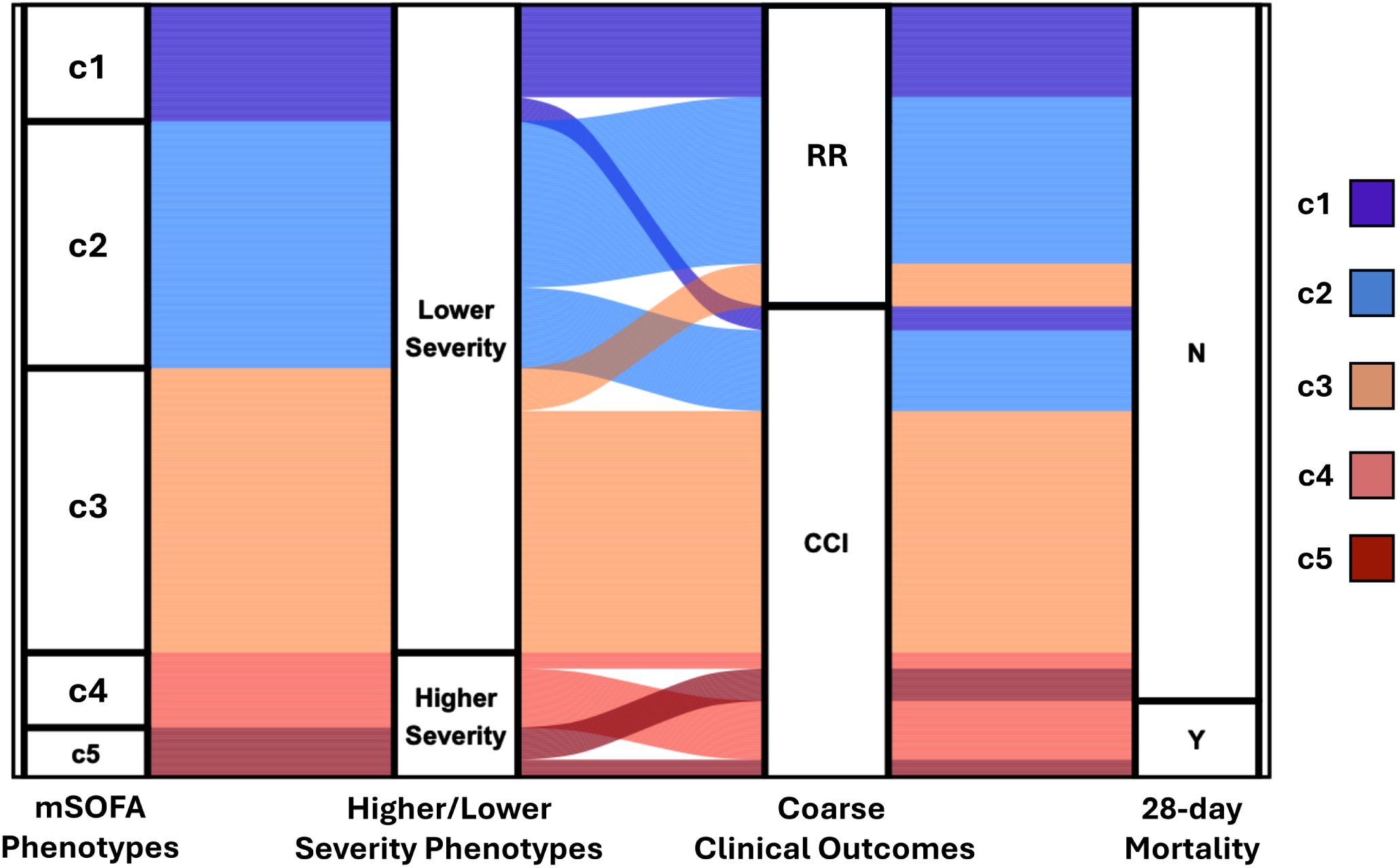
Phenotype relation to clinical outcome and mortality. Alluvial plot flow of mSOFA trajectory phenotype membership to coarse clinical outcome and 28-day mortality (n=69). Nodes include phenotype (c1-c5), higher/lower severity classification, coarse clinical outcome (rapid recovery, RR; chronic critical illness, CCI) and 28-day mortality. Cluster sizes: c1 (n=14), c2 (n=22), c3 (n=21), c4 (n=7), c5 (n=5).

### Organ dysfunction c1-c5 phenotype characteristics

The five mSOFA trajectory phenotypes also revealed unique patterns of organ system dysfunction, as visualized in Figure S3 (SDC4). Phenotype c1 represents a pattern of rapidly resolving cardiovascular and respiratory dysfunction. Phenotypes c2 and c3 reveal patterns of early multiple organ dysfunction (cardiac, respiratory & renal) with increasingly delayed trajectories of recovery. Phenotype c5 was one of fulminant and persistent multiple organ failure, including coagulopathy, liver dysfunction and renal failure. Phenotype c4 exhibited a unique pattern of persistent, moderate organ dysfunction with late fulminant renal failure. Additional phenotype characteristics can be appreciated in the individual organ system panels of Figure S3 (SDC4). Cardiovascular dysfunction (i.e., vasopressor intensity/duration) increases progressively across the 5 phenotypes. The c3, c4 and c5 phenotypes are characterized by prolonged respiratory dysfunction and ventilator dependence. Persistent coagulopathy and liver dysfunction are unique to the c5 phenotype.

Interestingly, the five mSOFA trajectory phenotypes show no significant differences across a wide range of demographics and clinical characteristics including sex, race, ethnicity, injury type/mechanism, and presence of major comorbidity (Figure 3; SDC4 Figures S4A/B/C). Additionally, injury severity score (ISS) and early physiologic parameters such as Emergency Department (ED) lowest systolic blood pressure, ED highest heart rate, and initially measured serum lactate level were not statistically different across all 5 phenotypes (Figure 3; SDC4 Figures S4A/B/C). However, we did identify significant differences in total resuscitation requirements (packed red blood cells, fresh frozen plasma, platelets), particularly in the c3 and c5 phenotypes, as well as in severity and duration of shock as measured by peak serum lactate through 24 hours (**Figure 3**). Additionally, the c4 phenotype was significantly older in age compared to the other phenotypes (**Figure 3**).

**Figure 3.**
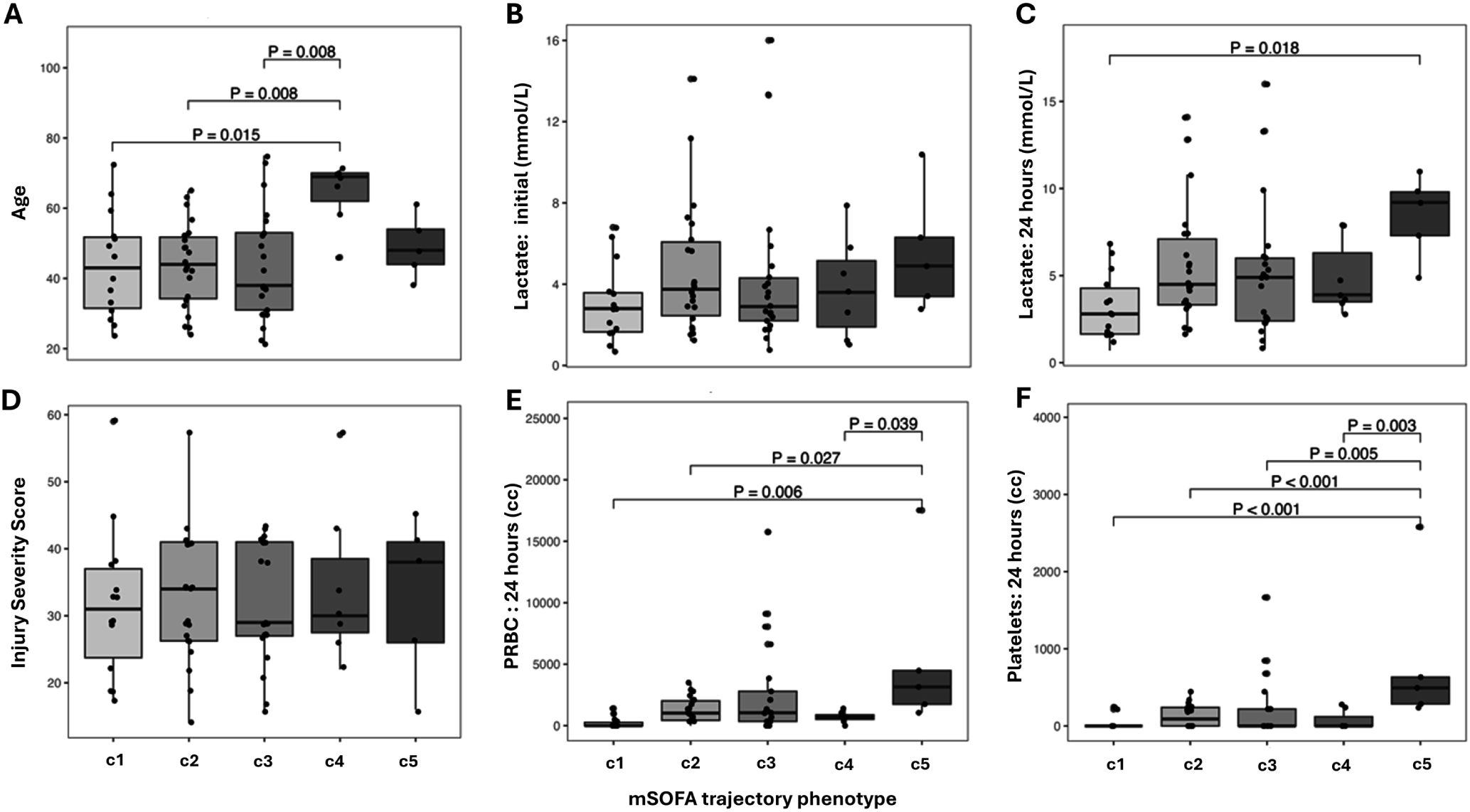
Clinical characteristics across phenotypes. Clinical characteristics compared across phenotypes. (A) age, (B) initial measured lactate, (C) peak lactate through 24 hours, (D) Injury Severity Score, (E) packed red blood cell volume transfused through 24 hours, and (F) platelet volume transfused through 24 hours. Significant across group and pairwise comparisons (p<0.05) are shown.

### Immune proteomic signatures across organ dysfunction phenotypes

Mixed effects and time-interaction modeling revealed significant differences in the peak abundance and longitudinal trajectories of specific plasma immune markers. Forty of the 92 measured plasma immune-related proteins demonstrated differences in longitudinal trajectory across the 5 mSOFA phenotypes (FDR<0.1; SDC3 Table S2). Figure S5 illustrates the top 15 significant marker trajectories that differ across phenotypes (SDC4), which represent mechanisms of adaptive lymphocyte regulation (CD5, CD8A, IL-15RA, TNFRSF9, SIRT2), epithelial-barrier immune signaling (IL-17C, IL-20RA), and tissue-injury/remodeling (ARTN, VEGFA, FGF-23, HGF, NT-3, OPG, FGF-5; SDC3 Table S1).

Given the fulminant course and early mortality of the c5 phenotype we selected for longitudinal immune marker trajectories distinguishing it from the lower severity phenotypes (c1, c2, c3). There were 3 immune markers (IL-17C, CXCL5, CXCL6) with trajectories differentiating c5 from c1/c2/c3 phenotypes. The c5 phenotype had significantly higher initial but then declining levels of IL-17C and CXCL6 compared to the low-severity phenotypes (**Figure 4**). In contrast, the low-severity phenotypes exhibited an increasing trajectory of CXCL5 compared to the c5 phenotype (**Figure 4**). The primary mechanisms represented by these immune markers are epithelial barrier inflammation and immunity (IL-17C), and neutrophil trafficking and activation (CXCL5, CXCL6; SDC3 Table S3).

**Figure 4.**
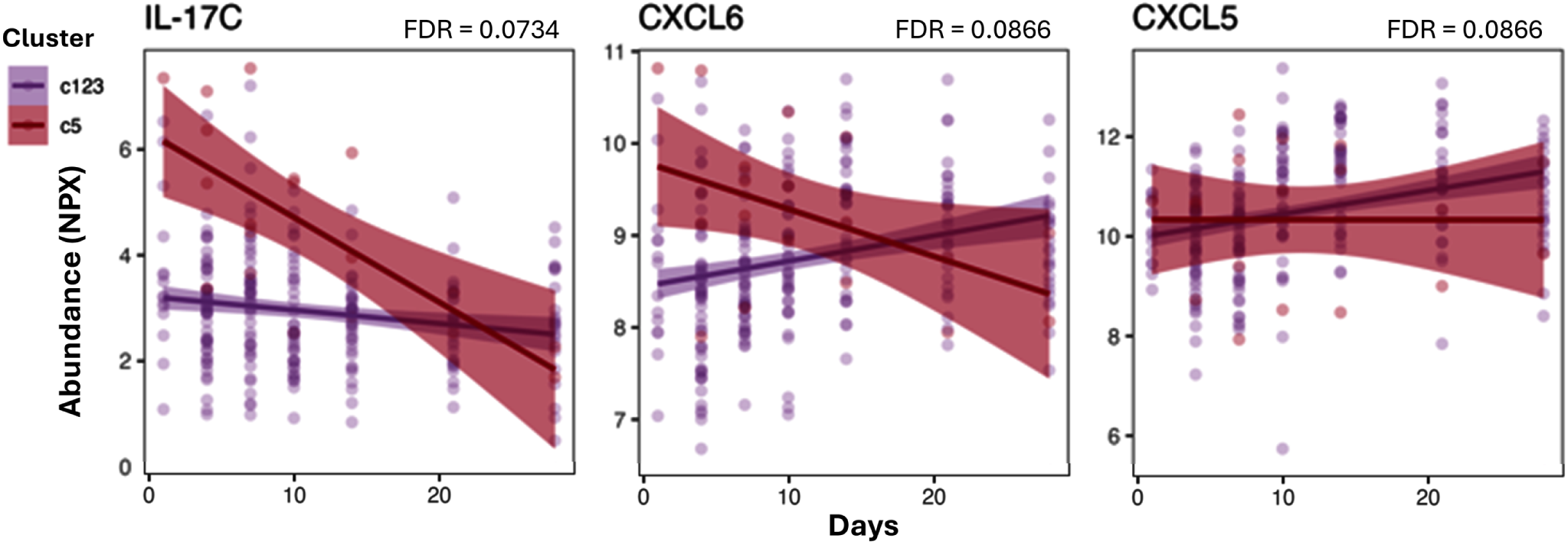
Distinguishing immune marker trajectories for phenotype c5. Plasma protein immune markers with significant difference in longitudinal trajectory (FDR<0.1; n=3) between phenotype c5 as compared to c1/c2/c3.

Additionally, considering the c3 phenotype was associated with a significantly higher proportion of patients developing chronic critical illness (**Figure 2**), we analyzed immune markers distinguishing it from the other lower-severity phenotypes (c1, c2). The analysis revealed 13 differentiating proteins (FDR<0.1) across the 3 lower-severity trajectories (SDC3 Table S4). Select differentiating marker trajectories for phenotype c3 are illustrated in Figure S6 (SDC 4). The c3 phenotype exhibited persistently rising trajectories of CXCL9, ARTN and TWEAK (Th1 recruitment/polarization and tissue inflammation/repair signaling; SDC3 Table S4). The c1 phenotype showed significantly increased trajectories of ADA, SIRT2 and STAMBP (innate inflammation resolution) and decreasing trajectories of CCL11, CST5 and IL-24 (leukocyte recruitment and reparative signaling functions; SDC3 Table S4).

Finally, given the unique clinical and organ dysfunction characteristics of the c4 phenotype, we selected for immune markers that differentiated this older-age phenotype from the remaining phenotypes (c1/2/3/5). We identified 29 immune-related protein markers with longitudinal trajectories differentiating the c4 group from the four other phenotypes including CD5, IL-15RA, FGF-23, CD8A and VEGFA (FDR<0.1, SDC3 Table S5). A select set of these markers is shown in **Figure 5**. The primary mechanisms represented by these differentiating markers in phenotype c4 include renal injury/repair (FGF-23, VEGFA), activated endothelium (CX3CL1), and lymphocyte adaptive immunity modulation (i.e., CD5, IL-15RA, IL-20RA; SDC3 Table S5).

**Figure 5.**
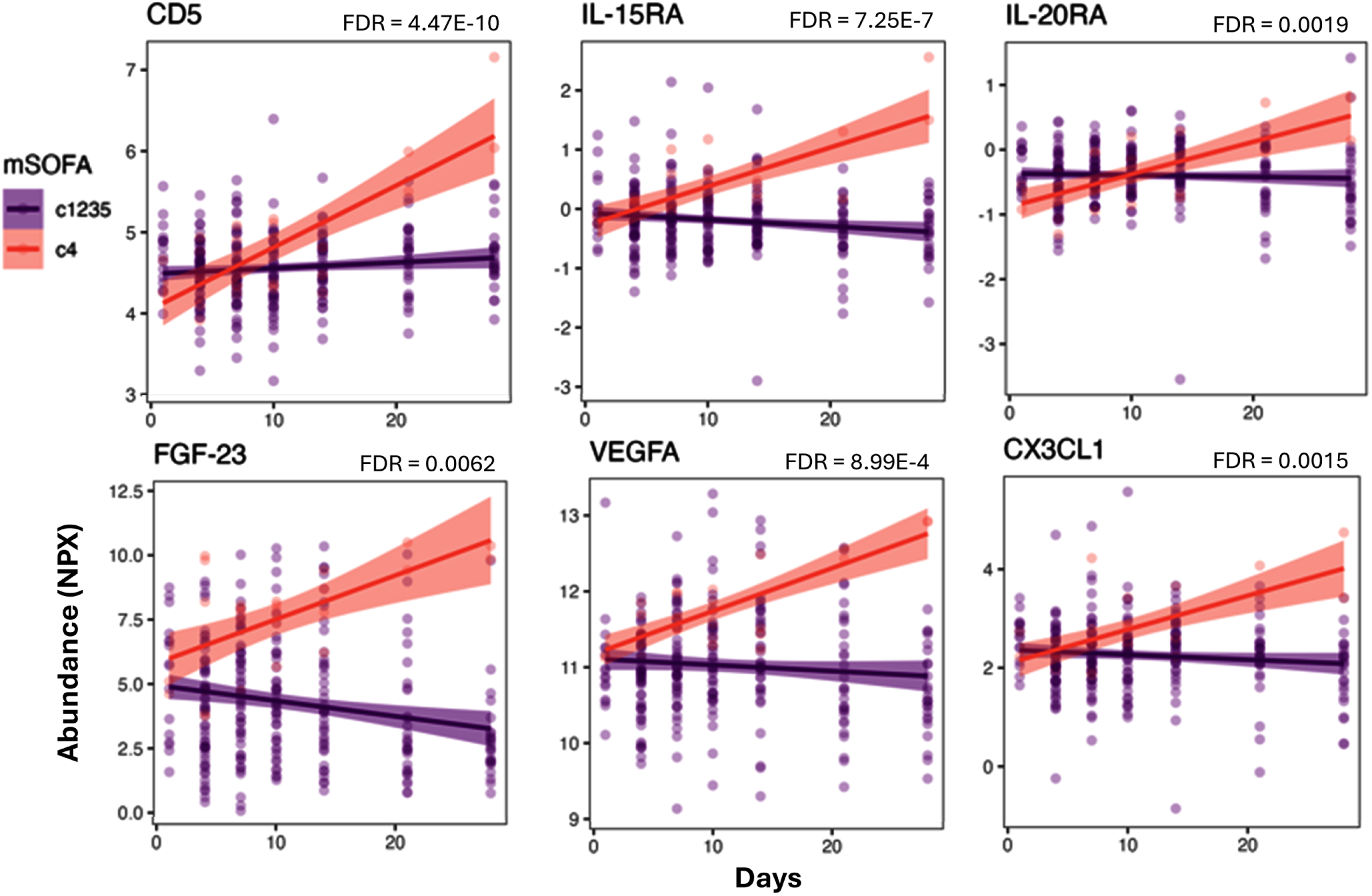
Distinguishing immune marker trajectories for phenotype c4. Six of the 29 plasma protein immune markers with significant difference in longitudinal trajectory (FDR<0.1) for phenotype c4 as compared to c1/c2/c3/c5. The full list of markers meeting FDR<0.1 (n=29) is listed in *SDC3 Table S5*.

## DISCUSSION

The heterogeneity of organ dysfunction after severe trauma is incompletely explained by clinical variables alone.(4, 20, 21) Without phenotype-level biologic characterization, immunomodulatory trials in critical illness risk repeating the failures seen when treatment is applied to an undifferentiated population. This study paired mSOFA score trajectory phenotyping with matched-timepoint immune proteomics in a cohort designed *a priori* for that pairing. We purposefully excluded the neurologic (GCS) subdomain from mSOFA and neurologically devastating TBI from the cohort itself, since GCS in this population primarily reflects TBI severity rather than the systemic host response to injury.(4, 22, 23) This two-layer design was intentional to isolate organ dysfunction driven by the post-injury host response rather than from primary neurologic injury.

We identified 5 distinct mSOFA trajectory phenotypes from a clinically homogeneous, severely injured, shock-enriched cohort. Forty of 92 measured plasma immune markers showed phenotype-differentiated longitudinal trajectories at an FDR<0.1, suggesting that these clinically derived trajectory groups are associated with distinct underlying immune biology rather than a single gradient of injury severity. Prior trajectory studies in broader trauma populations identified three to five organ dysfunction trajectories,(8, 9) suggesting that the present phenotypes represent further refinement within a high-risk subgroup rather than a competing classification.

The c5 phenotype, marked by most rapid/fulminant path to death, was differentiated by IL-17C, CXCL5, and CXCL6 trajectories, implicating epithelial barrier signaling and neutrophil trafficking. IL-17C drives epithelial barrier defense and neutrophil recruitment, acting as an autocrine immune amplifier.(24) IL-17C also has been shown to have an organ-specific role in pulmonary infection and ischemic renal injury(25–27), which is plausibly relevant given the shared hemorrhagic-shock physiology and high incidence of respiratory dysfunction and acute kidney injury. Of note, we have recently shown that CD4+ T helper 17 (Th17 cells) expansion and persistence are associated with the development of chronic critical illness (CCI) and recurrent nosocomial infections in severely injured trauma patients.(28) Thus, the Th17 axis warrants further investigation into its role in persistent, dysfunctional immunity after severe trauma. The divergent CXCL5/CXCL6 trajectories between c5 and the lower-severity phenotypes may also reflect contribution to compromised epithelial and tissue immunity through a progressive deterioration in ability to recruit neutrophils to sites of infection.(29, 30) Overall, this picture fits the known state of persistent dysfunctional immunity seen among critically injured trauma patients that survive to develop CCI, as their most common cause of death is recurrent multiple organ failure due to sepsis.(31)

The c3 phenotype, associated with the highest incidence of CCI, was distinguished by rising CXCL9, ARTN, and TWEAK. CXCL9 is IFNγ-induced and recruits CXCR3-expressing Th1 cells.(32, 33) TWEAK/Fn14 pro-inflammatory signaling is reparative when transient but becomes pathogenic with sustained activation(34), a pattern consistent with c3’s persistently rising TWEAK trajectory and its association with a chronic, unresolving clinical course. In contrast, c1 showed rising ADA, SIRT2, and STAMBP alongside declining CCL11, CST5, and IL-24. SIRT2 deacetylates and inhibits NLRP3 inflammasome activation(35, 36), offering one plausible basis for an inflammation-resolution interpretation in c1 that distinguishes it from c3’s sustained inflammatory-repair program. This persistent inflammatory signature in c3 is broadly consistent with the persistent inflammation component of the Persistent Inflammation, Immunosuppression and Catabolism Syndrome (PICS) as previously described in trauma patients(37, 38), though this proteomic panel does not assess the myeloid derived suppressor cell (MDSC)-driven immunosuppression arm central to PICS pathophysiology. We therefore do not claim c3 represents PICS, only that its inflammatory signature is consistent with one component of it.

The c4 phenotype, defined by older age, carried the largest differentiating signature (29 proteins), spanning renal injury/repair (FGF-23, VEGFA)(39–41), endothelial activation (CX3CL1)(42, 43), and adaptive lymphocyte modulation (CD5, IL-15RA, IL-20RA, CD8A).(44, 45) This is consistent with a distinct age-associated immune and renal-endothelial signature rather than being a demographic artifact of clustering. Additionally, this picture of persistently dysregulated signals that affect adaptive lymphocyte immunity is consistent with prior demonstration that PICS biomarker evidence of immunosuppression concentrates in older critically ill patients.(38, 46, 47)

Taken together, c5, c3, and c4 each present mechanistically distinct, non-overlapping marker signatures, consistent with separable immune biology rather than a single axis of increasing inflammatory magnitude. No single phenotype identified in this study completely recapitulates the full PICS triad. Phenotypes c3 and c4 each align with a different, partial component of PICS (persistent inflammation and an immunosuppression-adjacent adaptive signature, respectively), consistent with PICS as a composite syndrome that this analysis has resolved into distinguishable sub-signatures.

These findings have implications for the design of future therapeutic trials. Unrecognized biologic heterogeneity is a leading explanation for the repeated failure of mechanism-targeted therapies in critical illness. In sepsis, clinically derived phenotypes altered the apparent benefit or harm of treatment in simulated trials,(48) and in acute respiratory distress syndrome, subphenotypes responded differently to the same intervention.(49) The present data suggest a similar problem after severe injury. Phenotypes c4 and c5 shared a high risk of death, yet differed in organ dysfunction pattern, resuscitation burden, and immune signature, making it unlikely that a single mechanism-targeted therapy would benefit both. Conversely, c1 and c3 were indistinguishable at baseline but diverged in both trajectory and immune profile, suggesting that a targeted therapy whose effect would be diluted in the broader population could meaningfully reduce CCI in the c3 group. Once validated, organ dysfunction trajectory phenotypes may provide a framework for prognostic and predictive enrichment in future trials.

Several limitations need to be acknowledged. This is a single-center cohort of 69 patients delineating five trajectory phenotypes. Our presented cluster model metrics should be weighed directly against the risk of overfitting at this sample size, although high posterior classification probabilities indicate low assignment ambiguity, including for the smaller c4 and c5 subgroups. The deliberate exclusion of the neurologic subdomain from mSOFA potentially leaves residual variability from non-devastating head injury among included patients uncaptured by the organ dysfunction score. Additionally, it is highly unlikely that any single center/cohort study will fully delineate all generalizable disease phenotypes. Both confirmatory and complementary studies will be required to fully understand the scope and complexity of host response heterogeneity after severe trauma. Thus, all mechanism-phenotype associations are acknowledged as correlative and should be considered exploratory. Transfusion volume and shock severity were intentionally not adjusted for, as these variables lie on the causal pathway between injury and organ dysfunction; adjusting for such intermediates can remove the biology that defines a phenotype rather than confounding.(50) Other host and treatment factors with known immunologic effects, including sex, injury mechanism, and the type and magnitude of operative interventions, were not modeled as covariates. Although sex and injury mechanism did not differ significantly across phenotypes (SDC4 Figure S4B), the small size of some phenotypes limits the power to detect such differences. Finally, phenotype assignment was based on the full 28-day trajectory, and earlier identification and potential diagnostic biomarkers of these phenotypes will require further study.

Overall, we have shown here that there is significant underrecognized heterogeneity in organ dysfunction after severe trauma and hemorrhagic shock. Additionally, these post-trauma mSOFA phenotypes show evidence of unique biologic signatures that align with known mechanisms of adverse outcomes in critically ill surgical patients. These novel phenotypes and specific biologic signatures are hypothesis-generating rather than confirmatory. Their principal contribution is to reframe post-injury organ dysfunction trajectory as a biologically meaningful stratification variable, one that may ultimately support more targeted mechanistic intervention than current stratification that injury or shock severity can provide.

## DATA AVAILABILITY STATEMENT

De-identified participant-level data underlying this study will be made available to qualified investigators on reasonable request to the corresponding author, to support reproducibility and secondary analyses. These data comprise daily mSOFA scores and phenotype assignments, baseline demographic, injury, and resuscitation variables, and plasma Olink Target 96 Inflammation panel protein data (NPX values). Sharing will follow NIH data sharing policy, University of Washington Institutional Review Board approval, participant informed consent, and applicable privacy regulations. Methods and phenotype-specific immune marker results are provided in Supplemental Digital Content 2 to 4.

## SUPPLEMENTAL DIGITAL CONTENT

SDC 1. STROBE Checklist and Patient Flow Diagram.

SDC 2. Expanded Methods. Inclusion and exclusion criteria, informed consent, modified SOFA (mSOFA) scoring and imputation, trajectory model selection and classification diagnostics, statistical methods, and protein function annotation.

SDC 3. Supplemental Tables. Tables S1 to S5: plasma immune protein function/mechanism annotations and phenotype-specific immune marker analyses.

SDC 4. Supplemental Figures. Figures S1 to S6: individual mSOFA trajectories, hospital length of stay, organ system trajectories, clinical characteristics, and immune marker trajectories across phenotypes.

## Author Contributions

Conception and study design: HD, SCB. Analysis and interpretation of data: MC, QQ, NM, DB, KG, KS, KW, GOK, SCB. Drafting of the manuscript: SCB. Critical revision: HD, MC, QQ, NM, DB, KG, KS, KW, GOK, SCB.

This study was presented at the 85th annual meeting of the American Association for the Surgery of Trauma, September 18th, 2026, in Dallas, Texas. Acknowledgements: The authors would like to thank Keegan Stromberg, Sonja Katt and Laura Hennessy for their invaluable contribution to the successful execution of this study.

## Supporting information

SDC1 – STROBE checklist

SDC2 – Expanded Methods

SDC3 – Supplemental Tables

SDC4 – Supplemental Figures

## REFERENCES

1. Gentile LF, Cuenca AG, Efron PA, Ang D, Bihorac A, McKinley BA, et al. Persistent inflammation and immunosuppression: a common syndrome and new horizon for surgical intensive care. J Trauma Acute Care Surg. 2012;72(6):1491–1501.

2. Mira JC, Cuschieri J, Ozrazgat-Baslanti T, Wang Z, Ghita GL, Loftus TJ, et al. The epidemiology of chronic critical illness after severe traumatic injury at two level-one trauma centers. Crit Care Med. 2017;45(12):1989–1996.

3. Hawkins RB, Raymond SL, Stortz JA, Horiguchi H, Brakenridge SC, Gardner A, et al. Chronic critical illness and the persistent inflammation, immunosuppression, and catabolism syndrome. Front Immunol. 2018;9:1511.

4. Xiao W, Mindrinos MN, Seok J, Cuschieri J, Cuenca AG, Gao H, et al. A genomic storm in critically injured humans. J Exp Med. 2011;208(13):2581–2590.

5. Cabrera CP, Manson J, Shepherd JM, Torrance HD, Watson D, Longhi MP, et al. Signatures of inflammation and impending multiple organ dysfunction in the hyperacute phase of trauma: a prospective cohort study. PLoS Med. 2017;14(7):e1002352.

6. Chen T, Hughes J, Cornoy J, Chen W, Duerr R, Billiar T. Multi-modal single-cell sequencing reveals network transition in circulating monocytes that aligns with faster recovery in patients with trauma and favours a response to M-CSF. EBioMedicine. 2026;130:106368.

7. Brakenridge SC, Wang Z, Cox M, Raymond S, Hawkins R, Darden D, et al. Distinct immunologic endotypes are associated with clinical trajectory after severe blunt trauma and hemorrhagic shock. J Trauma Acute Care Surg. 2021;90(2):257–267.

8. Eriksson J, Nelson D, Holst A, Hellgren E, Friman O, Oldner A. Temporal patterns of organ dysfunction after severe trauma. Crit Care. 2021;25(1):165.

9. Cole E, Gillespie S, Vulliamy P, Brohi K; Organ Dysfunction in Trauma (ORDIT) study collaborators. Multiple organ dysfunction after trauma. Br J Surg. 2020;107(4):402–412.

10. Cohen MJ, Erickson CB, Lacroix IS, Debot M, Dzieciatkowska M, Sen S, et al. Multiomic analyses of longitudinal plasma samples identify thromboinflammation endotypes and trajectories in patients with trauma. Sci Transl Med. 2026;18(836):eadw5223.

11. Vincent JL, Moreno R, Takala J, Willatts S, De Mendonça A, Bruining H, et al. The SOFA (Sepsis-related Organ Failure Assessment) score to describe organ dysfunction/failure. Intensive Care Med. 1996;22(7):707–710.

12. Zygun DA, Kortbeek JB, Fick GH, Laupland KB, Doig CJ. Non-neurologic organ dysfunction in severe traumatic brain injury. Crit Care Med. 2005;33(3):654–660.

13. Gordon AC, Perkins GD, Singer M, McAuley DF, Orme RML, Santhakumaran S, et al. Levosimendan for the prevention of acute organ dysfunction in sepsis. N Engl J Med. 2016;375(17):1638–1648.

14. Moreno R, Rhodes A, Piquilloud L, Hernandez G, Takala J, Gershengorn HB, et al. The Sequential Organ Failure Assessment (SOFA) score: has the time come for an update? Crit Care. 2023;27(1):15.

15. Alrawashdeh M, Klompas M, Rhee C. The impact of common variations in Sequential Organ Failure Assessment score calculation on sepsis measurement using Sepsis-3 criteria: a retrospective analysis using electronic health record data. Crit Care Med. 2024;52(9):1380–1390.

16. Dill-McFarland KA, Mitchell K, Batchu S, Segnitz RM, Benson B, Janczyk T, et al. Kimma: flexible linear mixed effects modeling with kinship covariance for RNA-seq data. Bioinformatics. 2023;39(5):btad279.

17. Benjamini Y, Hochberg Y. Controlling the false discovery rate: a practical and powerful approach to multiple testing. J Roy Stat Soc B. 1995;57(1):289–300.

18. Karabatsiakis A, Hamuni G, Wilker S, Kolassa S, Renu D, Kadereit S, et al. Metabolite profiling in posttraumatic stress disorder. J Mol Psychiatry. 2015;3(1):2.

19. Reiner A, Yekutieli D, Benjamini Y. Identifying differentially expressed genes using false discovery rate controlling procedures. Bioinformatics. 2003;19(3):368–375.

20. Xu Z, Mao C, Su C, Zhang H, Siempos I, Torres LK, et al. Sepsis subphenotyping based on organ dysfunction trajectory. Crit Care. 2022;26(1):197.

21. Yang Z, Le T, Simovic MO, Liu B, Fraker TL, Cancio TS, et al. Traumatized triad of complementopathy, endotheliopathy, and coagulopathy – impact on clinical outcomes in severe polytrauma patients. Front Immunol. 2022;13:991048.

22. Krishnamoorthy V, Temkin N, Barber J, Foreman B, Komisarow J, Gepts CM, et al. Association of early multiple organ dysfunction with clinical and functional outcomes over the year following traumatic brain injury: a TRACK-TBI study. Crit Care Med. 2021;49(10):1769–1778.

23. Vasilevskis EE, Pandharipande PP, Graves AJ, Shintani A, Tsuruta R, Ely EW, et al. Validity of a modified Sequential Organ Failure Assessment score using the Richmond Agitation-Sedation Scale. Crit Care Med. 2016;44(1):138–146.

24. Ramirez-Carrozzi V, Sambandam A, Luis E, Lin Z, Jeet S, Lesch J, et al. IL-17C regulates the innate immune function of epithelial cells in an autocrine manner. Nat Immunol. 2011;12(12):1159–1166.

25. Zhang F, Yin J, Liu L, Liu S, Zhang G, Kong Y, et al. IL-17C neutralization protects the kidney against acute injury and chronic injury. EBioMedicine. 2023;92:104607.

26. Wolf L, Sapich S, Honecker A, Jungnickel C, Astaburuaga-García R, Pinto AL, et al. IL-17A-mediated expression of epithelial IL-17C promotes inflammation during acute Pseudomonas aeruginosa pneumonia. Am J Physiol Lung Cell Mol Physiol. 2016;311(5):L1015–L1022.

27. Wang F, Yin J, Lin Y, Zhang F, Liu X, Zhang G, et al. IL-17C has a pathogenic role in kidney ischemia/reperfusion injury. Kidney Int. 2020;97(6):1219–1229.

28. Kim CY, Ding H, Mirabadi N, Luo S, Scherler K, Walters KA, et al. Distinct immune trajectories after severe trauma identify a Th17-biased immunophenotype in chronic critical illness. J Immunol. 2026;215(6):vkag125.

29. Madalli S, Beyrau M, Whiteford JR, Duchene J, Singh Nandhra I, Patel NS, et al. Sex-specific regulation of chemokine Cxcl5/6 controls neutrophil recruitment and tissue injury in acute inflammatory states. Biol Sex Differ. 2015;6:27.

30. Linge HM, Collin M, Nordenfelt P, Mörgelin M, Malmsten M, Egesten A. The human CXC chemokine granulocyte chemotactic protein 2 (GCP-2)/CXCL6 possesses membrane-disrupting properties and is antibacterial. Antimicrob Agents Chemother. 2008;52(7):2599–2607.

31. Brakenridge SC, Efron PA, Cox MC, Stortz JA, Hawkins RB, Ghita G, et al. Current epidemiology of surgical sepsis: discordance between inpatient mortality and 1-year outcomes. Ann Surg. 2019;270(3):502–510.

32. Tokunaga R, Zhang W, Naseem M, Puccini A, Berger MD, Soni S, et al. CXCL9, CXCL10, CXCL11/CXCR3 axis for immune activation – a target for novel cancer therapy. Cancer Treat Rev. 2018;63:40–47.

33. Manicone AM, Burkhart KM, Lu B, Gerard NP, Gerard C, Wang X, et al. CXCR3 ligands contribute to Th1-induced inflammation but not to homing of Th1 cells into the lung. Exp Lung Res. 2008;34(7):391–407.

34. Winkles JA. The TWEAK-Fn14 cytokine-receptor axis: discovery, biology and therapeutic targeting. Nat Rev Drug Discov. 2008;7(5):411–425.

35. He M, Chiang HH, Luo H, Zheng Z, Qiao Q, Wang L, et al. An acetylation switch of the NLRP3 inflammasome regulates aging-associated chronic inflammation and insulin resistance. Cell Metab. 2020;31(3):580–591.

36. Misawa T, Takahama M, Kozaki T, Lee H, Zou J, Saitoh T, et al. Microtubule-driven spatial arrangement of mitochondria promotes activation of the NLRP3 inflammasome. Nat Immunol. 2013;14(5):454–460.

37. Vanzant EL, Lopez CM, Ozrazgat-Baslanti T, Ungaro R, Davis R, Cuenca AG, et al. Persistent inflammation, immunosuppression, and catabolism syndrome after severe blunt trauma. J Trauma Acute Care Surg. 2014;76(1):21–29.

38. Darden DB, Brakenridge SC, Efron PA, Ghita GL, Brumback B, Mohr AM, et al. Biomarker evidence of the persistent inflammation, immunosuppression and catabolism syndrome (PICS) in chronic critical illness (CCI) after surgical sepsis. Ann Surg Open. 2021;274(4):664–673.

39. Leaf DE, Jacob KA, Srivastava A, Chen ME, Christov M, Jüppner H, et al. Fibroblast growth factor 23 levels associate with AKI and death in critical illness. J Am Soc Nephrol. 2017;28(6):1877–1885.

40. Ren J, Liu K, Wu B, Wang F, Cheng M, Ye X, et al. Divergent actions of renal tubular and endothelial type 1 IL-1 receptor signaling in toxin-induced AKI. J Am Soc Nephrol. 2023;34(10):1629–1646.

41. Kang DH, Hughes J, Mazzali M, Schreiner GF, Johnson RJ. Impaired angiogenesis in the remnant kidney model: II. Vascular endothelial growth factor administration reduces renal fibrosis and stabilizes renal function. J Am Soc Nephrol. 2001;12(7):1448–1457.

42. Bazan JF, Bacon KB, Hardiman G, Wang W, Soo K, Rossi D, et al. A new class of membrane-bound chemokine with a CX3C motif. Nature. 1997;385(6617):640–644.

43. Rodriguez C, Chocarro L, Echaide M, Ausin K, Escors D, Kochan G. Fractalkine in health and disease. Int J Mol Sci. 2024;25(15):8007.

44. Hohlstein P, Gussen H, Bartneck M, Warzecha KT, Roderburg C, Buendgens L, et al. Prognostic relevance of altered lymphocyte subpopulations in critical illness and sepsis. J Clin Med. 2019;8(3):353.

45. Barrios EL, Balzano-Nogueira L, Polcz VE, Loftus TJ, Ozrazgat-Baslanti T, Sarosi GA, et al. Unique lymphocyte transcriptomic profiles in septic patients with chronic critical illness. Front Immunol. 2024;15:1478471.

46. Mankowski RT, Anton SD, Ghita GL, Brumback B, Darden DB, Bihorac A, et al. Older adults demonstrate biomarker evidence of the persistent inflammation, immunosuppression, and catabolism syndrome (PICS) after sepsis. J Gerontol A Biol Sci Med Sci. 2022;77(1):188–196.

47. Sathe NA, Bhatraju PK, Mikacenic C, Rue TC, Chai XY, Wurfel MM, et al. Relationships between age, soluble triggering receptor expressed on myeloid cells-1 (sTREM-1), and mortality among critically ill adults: a cohort study. Shock. 2021;57(2):205–211.

48. Seymour CW, Kennedy JN, Wang S, Chang CH, Elliott CF, Xu Z, et al. Derivation, validation, and potential treatment implications of novel clinical phenotypes for sepsis. JAMA. 2019;321(20):2003–2017.

49. Calfee CS, Delucchi K, Parsons PE, Thompson BT, Ware LB, Matthay MA; NHLBI ARDS Network. Subphenotypes in acute respiratory distress syndrome: latent class analysis of data from two randomised controlled trials. Lancet Respir Med. 2014;2(8):611–620.

50. Schisterman EF, Cole SR, Platt RW. Overadjustment bias and unnecessary adjustment in epidemiologic studies. Epidemiology. 2009;20(4):488–495.

