## Supplementary material for "Multiple Organ Dysfunction Trajectory Phenotypes After Severe Trauma Are Associated With Distinct Immune Profiles and Outcomes": SDC1 – STROBE checklist

**Supplemental Digital Content 1. STROBE Checklist and Patient Flow Diagram**

**STROBE Patient Flow Diagram**

Patient flow diagram for the prospective trauma cohort. Of 544 patients assessed for eligibility, 470 did not meet inclusion/exclusion criteria, leaving 74 eligible patients approached for enrollment. Five patients refused participation or were excluded after enrollment (samples destroyed and not included in the analytic cohort), yielding a final analytic cohort of 69 patients. Seventeen of the 69 enrolled/analyzed patients (24.6%) were enrolled under an IRB-approved conditional waiver of informed consent.

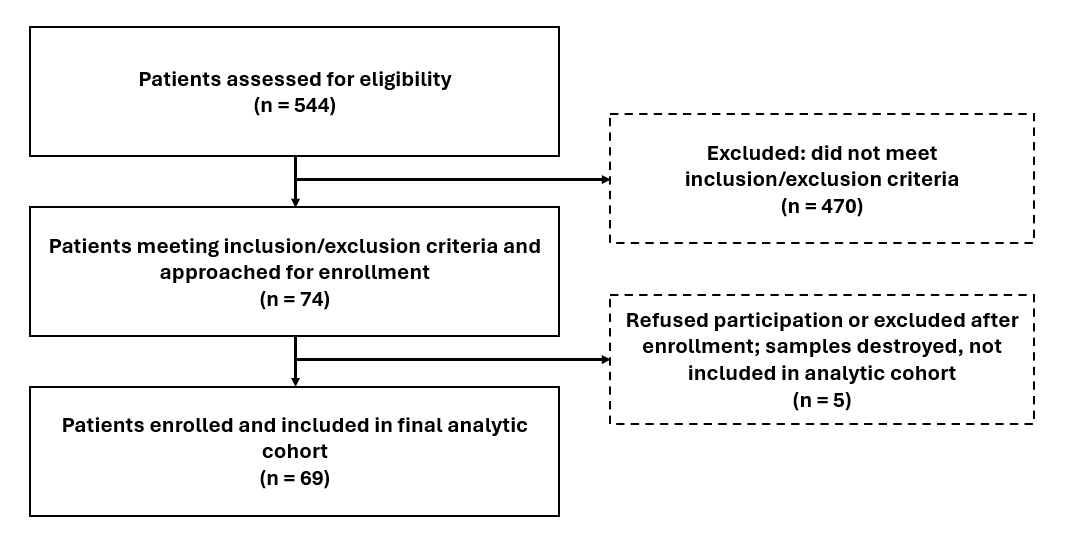

**STROBE Checklist for Cohort Studies**

| **Item** | **STROBE Topic** | **Status** | **Manuscript Location** |
| --- | --- | --- | --- |
| 1a | Title indicates study design | Reported | Abstract (Methods: prospective, longitudinal, observational cohort study) |
| 1b | Abstract: informative summary | Reported | Abstract |
| 2 | Background/rationale | Reported | Background (p. 1-2) |
| 3 | Objectives/hypotheses | Reported | Background, hypotheses (p. 2) |
| 4 | Study design | Reported | Methods, Study design (p. 2); SDC1 Expanded Methods |
| 5 | Setting, locations, dates | Reported | Methods, Study design (p. 2); Results, enrollment dates (p. 6); SDC1 Expanded Methods |
| 6a | Eligibility criteria; selection methods | Reported | Methods, Study design: inclusion/exclusion criteria and PI adjudication (p. 2); SDC1 Expanded Methods 1.1-1.2 |
| 6b | Matching criteria (if applicable) | N/A | Not a matched-cohort design |
| 7 | Variables: outcomes, predictors, confounders | Reported | Methods, Clinical data collection and outcome definitions (p. 4); Methods, GBTM and statistical analyses (p. 5) |
| 8 | Data sources/measurement | Reported | Methods, Biospecimen collection and mSOFA assessment (p. 3-4); SDC1 Expanded Methods 3.1-3.4 |
| 9 | Bias | Reported | Methods: prospective PI-adjudicated enrollment (p. 2), exclusion of devastating TBI and GCS subdomain (p. 2-3), a priori imputation (p. 4); Discussion, Limitations (p. 14) |
| 10 | Study size/justification | Partial | Results: all eligible patients enrolled over a fixed enrollment period, March 2022 to December 2024 (p. 6); exploratory design without a priori sample size calculation; sample size acknowledged in Discussion, Limitations (p. 14) |
| 11 | Quantitative variables: handling | Reported | Methods, mSOFA scoring and GBTM (p. 3-5); SDC1 Expanded Methods 3.1-4.1 |
| 12a | Statistical methods, incl. confounders | Reported | Methods, GBTM and statistical analyses (p. 5); Discussion, Limitations: rationale for not adjusting for causal-pathway variables (p. 14); SDC1 Expanded Methods 4.1-4.3 |
| 12b | Subgroup/interaction analyses | Reported | Methods, mixed-effects phenotype-by-time interaction models (p. 5); Results (p. 9-10) |
| 12c | Missing data handling | Reported | Methods, mSOFA imputation (p. 4) and proteomic limit-of-detection exclusion (p. 3); SDC1 Expanded Methods 3.3-3.4 |
| 12d | Loss to follow-up | Reported | Methods: patients followed until hospital discharge or death; no loss to follow-up during hospitalization (p. 4) |
| 12e | Sensitivity analyses | Partial | SDC1 Expanded Methods 4.1: comparison of alternative polynomial orders and cluster numbers, cross-validation of cluster assignment (latrend package), and classification certainty (APPA); Methods (p. 5) |
| 13a | Participant numbers at each stage | Reported | SDC2 Flow Diagram; Results, qualifying criteria and analytic cohort (p. 6) |
| 13b | Reasons for non-participation | Reported | SDC2 Flow Diagram |
| 13c | Flow diagram | Reported | SDC2 Flow Diagram |
| 14a | Descriptive data (characteristics) | Reported | Results (p. 6, 8); Table 1; Figure 3; SDC4 Figures S4A-C |
| 14b | Missing data per variable | Partial | SDC1 Expanded Methods 3.3-3.4 (mSOFA imputation); missing data not itemized per variable |
| 14c | Follow-up time/summary | Reported | Methods: followed through 28 days, death, or hospital discharge (p. 4-5) |
| 15 | Outcome data (event counts) | Reported | Results: RR/CCI and 28-day mortality by phenotype, including early deaths (p. 6-7); Figure 2 |
| 16a | Main results (estimates + precision) | Reported | Results (p. 6-10); Figures 1-5; SDC3 Tables S2-S5 (FDR-adjusted estimates); SDC4 Supplemental Figures |
| 16b | Category boundaries | N/A | No categorized continuous exposure |
| 16c | Absolute risk (if relevant) | Reported | Results: mortality and CCI proportions by phenotype (p. 6-7) |
| 17 | Other analyses (subgroup/sensitivity) | Reported | Results: c5 vs c1-c3, c3 vs c1-c2, c4 vs others (p. 9-10); SDC3 Tables S3-S5; SDC4 Figures S5-S6 |
| 18 | Key results summary | Reported | Discussion (p. 11) |
| 19 | Limitations | Reported | Discussion, Limitations (p. 14) |
| 20 | Interpretation | Reported | Discussion (p. 11-15) |
| 21 | Generalizability | Reported | Discussion, Limitations: single-center cohort; confirmatory and complementary studies required (p. 14) |
| 22 | Funding | Partial | Title page (funding sources). The funders had no role in study design, data collection, analysis, interpretation, or manuscript preparation |
