## Supplementary material for "Multiple Organ Dysfunction Trajectory Phenotypes After Severe Trauma Are Associated With Distinct Immune Profiles and Outcomes": SDC2 – Expanded Methods

**Supplemental Digital Content 2 (SDC2) – Expanded Methods**

***1.1. Inclusion Criteria***

Patients admitted to the Trauma ICU service at [*blinded*] were eligible for inclusion if they met either of the following criteria:

- Injury Severity Score (ISS) ≥25, and/or
- Evidence of hemorrhagic shock, defined as systolic blood pressure <90 mmHg or lactate >2 mmol/L on presentation.

Potential enrollees were prospectively adjudicated by the primary investigator (PI), including preliminary ISS score calculation (MDCalc ISS calculator, https://www.mdcalc.com/), and confirmation of meeting shock criteria. Final ISS scores were collected after completion by the institutional trauma registry.

***1.2. Exclusion Criteria***

Patients meeting any of the following criteria were excluded:

- Not expected to survive >48 hours from injury
- Investigator-adjudicated, neurologically devastating traumatic brain injury
- Prisoners or pregnant patients
- Chronic corticosteroid use, immunosuppressive therapy, or prior bone marrow transplantation
- Severe or confounding medical comorbidity, including: pre-existing end-stage renal disease; severe congestive heart failure (NYHA Class IV); chronic liver disease with MELD score ≥15; HIV infection with CD4+ count <200 cells/mm³; or active/symptomatic COVID-19 infection

The neurologically devastating TBI exclusion is applied in addition to removal of the Glasgow Coma Scale/central nervous system subdomain from the modified SOFA (mSOFA) scoring schema (see *mSOFA Organ Dysfunction Assessment and Scoring* below), representing a two-layer strategy to reduce confounding of systemic organ dysfunction by primary neurologic injury. This exclusion was determined prior to enrollment by adjudication of initial Neurosurgery consultation documentation by the primary investigator.

***2.1. Regulatory Approval and Informed Consent***

This study was approved by the [*blinded*] Institutional Review Board (IRB; protocol #00006241]) in full compliance with the Declaration of Helsinki and was designated “minimal risk” human subjects research. An IRB-approved study information sheet was provided, and informed consent was obtained from the patient or their legally authorized representative (i.e., “proxy”) if the patient was incapacitated, at any point during the hospitalization. If the patient never regained decisional capacity and no proxy could be identified or contacted, the IRB has granted a conditional waiver of informed consent for continued study participation. This conditional waiver policy is intended to permit initiation of research procedures deemed minimal risk in this difficult-to-consent, vulnerable population, balancing the principles of research social value, justice, equity, and autonomy against the practical constraints of obtaining timely consent after severe injury.

***3.1. mSOFA Organ Dysfunction Assessment and Scoring***

Post-injury organ dysfunction was quantified using a trauma-relevant modified Sequential Organ Failure Assessment (mSOFA) score, based on the standard SOFA score criteria of Vincent et al. (Intensive Care Med 1996), with two modifications (**Expanded Methods Table 1**).

First, the Glasgow Coma Scale (GCS)/central nervous system organ subscore was excluded from calculation of the total mSOFA score. This exclusion removes the confounding contribution of primary traumatic brain injury to a measure intended to quantify systemic, immune-driven multiple organ dysfunction, consistent with prior reports (Xiao et al., J Exp Med 2011; Zygun et al., J Crit Care 2003; Gordon et al., N Engl J Med 2016).

Second, two organ subscores were modified to reflect contemporary critical care support practices, as suggested in recent literature (Moreno, Vincent et al., Crit Care 2023):

- Respiratory: “with respiratory support” was expanded beyond invasive mechanical ventilation to include high-flow nasal cannula (HFNC) and non-invasive positive pressure ventilation (NIPPV/BiPAP). The SpO2/FiO2 ratio was substituted when PaO2/FiO2 was not calculable on a given day (see Respiratory Score Strategy, below).
- Cardiovascular: fixed-dose supplemental vasopressin use (>0.01 U/min) was assigned a minimum Cardiovascular subscore of 3, reflecting its increasingly common use in trauma patients with hemorrhagic shock (Sims et al., JAMA Surg 2019).

**Expanded Methods Table M1. mSOFA scoring strategy**

| **Organ System** | **Parameter, units** | **0** | **1** | **2** | **3** | **4** |
| --- | --- | --- | --- | --- | --- | --- |
| Respiratory | PaO2/FiO2, mm Hg | ≥400 | 300–399 | 200–299 | 100–199 with respiratory support¹ | <100 with respiratory support¹ |
|  | SpO2/FiO2 ratio² | ≥512 | 357–511 | 214–356 | 89–213 with respiratory support | <89 with respiratory support |
| Coagulation | Platelets, ×10³/mm³ | ≥150 | 100–149 | 50–99 | 20–49 | <20 |
| Hepatic | Bilirubin, mg/dL | <1.2 | 1.2–1.9 | 2.0–5.9 | 6.0–11.9 | >12.0 |
| Cardiovascular | MAP, vasopressor dose (mcg/kg·min) | MAP ≥70 mmHg | MAP <70 mmHg | Dopamine <5 or dobutamine (any dose) | Dopamine 5.1–15, epinephrine ≤0.1, norepinephrine ≤0.1, or vasopressin >0.01 U/min | Dopamine >15, epinephrine >0.1, or norepinephrine >0.1 |
| Renal | Creatinine, mg/dL; UOP, mL/day | <1.2 | 1.2–1.9 | 2.0–3.4 | 3.5–4.9 or UOP <500 | >5.0 or UOP <200 |

¹“With respiratory support” includes high-flow nasal cannula oxygen, non-invasive positive pressure ventilation, and mechanical ventilation via endotracheal tube or tracheostomy.

²SpO2/FiO2 ratio was substituted when the PaO2/FiO2 value was not calculable on a given calendar day (see Respiratory Score Strategy, below).

***3.2. mSOFA General Scoring Rules***

- For each organ system, a subscore of 4 indicates the worst derangement and 0 indicates normal function.
- The Daily Organ System Score reflects the worst (highest scoring) value recorded for that organ system on a given calendar day (e.g., lowest PaO2/FiO2 or SpO2/FiO2, lowest platelet count, highest bilirubin, highest creatinine or lowest urine output).
- The Cardiovascular subscore follows a hierarchical schema: when a patient meets criteria for more than one non-mutually exclusive cardiovascular category on the same day, the higher (worse) score is assigned.
- The Daily mSOFA Score is the sum of the six Daily Organ System Scores (Respiratory, Coagulation, Hepatic, Cardiovascular, Renal; GCS/CNS excluded).
- All mSOFA components were derived from ICU clinical flowsheet and laboratory data within the institutional electronic medical record (EMR).

***3.3. mSOFA Imputation Strategy***

Missing values for calculation of the Daily Organ System Score were handled using a Last Observation Carried Forward (LOCF) strategy (Alrawashdeh et al., Crit Care Med 2024):

- If a component was missing on a given day, the value was carried forward from the most recent prior observation, regardless of the interval elapsed.
- If values were missing on the first calendar day of the encounter (including time in the emergency department), values were carried backward from the next calendar day; if the next calendar day was also missing, a score of 0 was assigned.
- For the Hepatic subscore specifically, given the infrequency of routine bilirubin measurement in this population, bilirubin was considered normal (≤1.2 mg/dL) when missing, unless and until an abnormal value (>1.2 mg/dL) was encountered. Once an abnormal bilirubin value occurred, it was carried forward until a normal value was measured, or until discharge/death.
- For patients who died during hospitalization, an mSOFA value of 20 was appended to the terminal timepoint; for patients discharged from the ICU/hospital, a value of −1 was appended, consistent with conventions for trajectory modeling of terminal clinical states.

***3.4. mSOFA Respiratory Score Strategy***

Respiratory subscore assignment followed a hierarchical decision rule (adapted from Alrawashdeh et al., Crit Care Med 2024):

- If a simultaneous (same-hour) PaO2 and FiO2 were available on a given calendar day, these were used directly to calculate the PaO2/FiO2 ratio.
- If FiO2 was not simultaneous with PaO2, FiO2 within ±1 hour was used; if unavailable, matching preceding and following FiO2 values within a ±3-hour window were used; if still unavailable, FiO2 was estimated from the recorded oxygen delivery device (see FiO2 conversion table below).
- If no PaO2 value was available on a given day, the SpO2/FiO2 (S/F) ratio was substituted. Simultaneous SpO2/FiO2 pairs (within ±10 minutes) were preferred; if unavailable, FiO2 within ±1 hour of the SpO2 was used; if still unavailable, matching preceding and following FiO2 values within a ±3-hour window were used. The lowest S/F ratio for the day was used to assign the daily subscore.

FiO2 was estimated from non-invasive oxygen delivery devices using the following conversion table when a directly measured FiO2 was unavailable:

| **Device** | **FiO2 conversion** |
| --- | --- |
| Room air | 0.21 |
| Nasal cannula 1 L | 0.24 |
| Nasal cannula 2 L | 0.27 |
| Nasal cannula 3 L | 0.30 |
| Nasal cannula 4 L | 0.33 |
| Nasal cannula 5 L | 0.36 |
| Nasal cannula 6 L | 0.39 |
| Nasal cannula 7 L | 0.42 |
| Nasal cannula 8 L | 0.45 |
| Face mask 5 L | 0.40 |
| Face mask 6–7 L | 0.50 |
| Face mask 7–8 L | 0.60 |
| Face mask, flow not specified | 0.40 |
| Non-rebreather 6 L | 0.60 |
| Non-rebreather 7 L | 0.70 |
| Non-rebreather 8 L | 0.80 |
| Non-rebreather 9 L | 0.90 |
| Non-rebreather, flow not specified | 0.60 |
| High-flow nasal cannula, BiPAP, or mechanical ventilation | Use direct FiO2 measurement |

***4.1. Cluster mSOFA Phenotype Development***

Latent class trajectory modeling was performed on SOFA scores over the first 28 days of hospitalization. In the case of death, a SOFA value of 20 was appended to the patient’s time scores, and in the case of discharge, a SOFA value of -1 was appended. Clustering was performed using the lcmm package (v2.2.2)^1^ in R v4.6.0^2^. Akaike Information Criterion (AIC)^3^, Bayesian Information Criterion (BIC)^4^, and log likelihood^5^ metrics aided in the choice of a second order polynomial model [SOFA ~ day + I(day^2)] with five outcome clusters (**Expanded Methods Figure M1**).

**Expanded Methods Figure M1. Akaike Information Criterion (AIC), Bayesian Information Criterion (BIC), and log likelihood metrics for c1-c4 longitudinal mSOFA cluster phenotypes.**

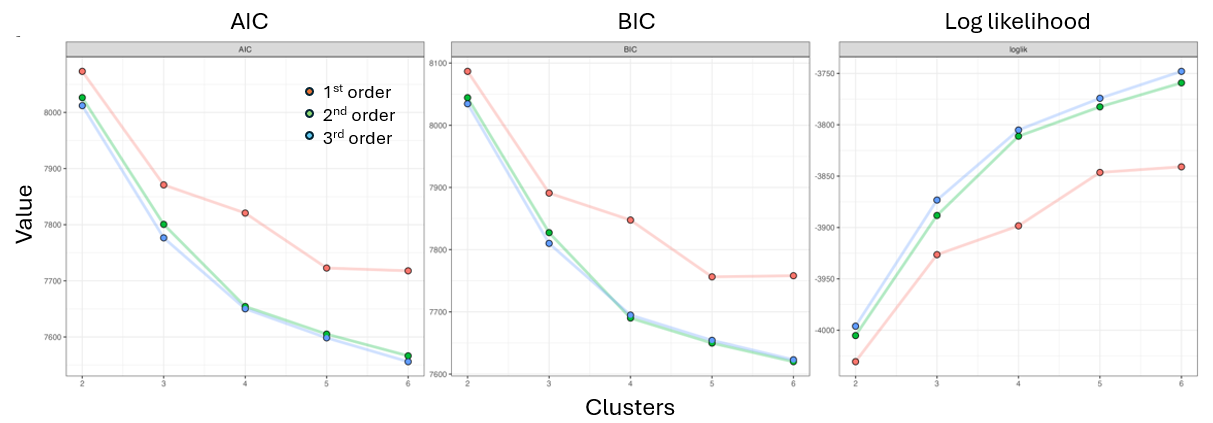

Model selection was performed using the "hlme" function within the lcmm package, comparing candidate polynomial orders (linear, quadratic, cubic) and cluster solutions (k=2 through k=6) based on AIC, BIC, and log-likelihood (**Expanded Methods Table 2**). As a cross-validation step, cluster assignments were compared against those generated using the latrend package. Cluster membership was highly concordant between the two methods, corroborated by comparable rapid recovery/chronic critical illness (RR/CCI) and 28-day mortality (M28) alignment across the 5- and 6-cluster solutions. Although 6-cluster solutions showed marginally improved fit statistics relative to 5-cluster solutions, the resulting subgroup sizes at 6 clusters were judged too small to support stable, clinically interpretable phenotypes. Because second- and third-order polynomial solutions produced highly similar fit and cluster structure, the second-order (quadratic) polynomial was selected for parsimony. The final model therefore consisted of a second-order polynomial, 5-cluster solution (c1-c5) fit using hlme, which was used for all subsequent phenotype-immune proteomic analyses.

**Expanded Methods Table M2. Group-based trajectory model fit statistics across candidate polynomial orders and cluster solutions.**

| **Polynomial order** | **Clusters (k)** | **AIC** | **BIC** | **Log-likelihood** |
| --- | --- | --- | --- | --- |
| ***A. Linear polynomial (day)*** | | | | |
|  | 2 | 8073.2 | 8086.6 | -4030.6 |
|  | 3 | 7870.9 | 7891.0 | -3926.5 |
|  | 4 | 7820.7 | 7847.6 | -3898.4 |
|  | 5 | 7722.8 | 7756.3 | -3846.4 |
|  | 6 | 7717.9 | 7758.1 | -3840.9 |
| ***B. Quadratic polynomial (day, day²)*** | | | | |
|  | 2 | 8026.4 | 8044.3 | -4005.2 |
|  | 3 | 7800.6 | 7827.4 | -3888.3 |
|  | 4 | 7654.3 | 7690.0 | -3811.1 |
| *Selected model →* | **5** | **7605.1** | **7649.8** | **-3782.5** |
|  | 6 | 7566.3 | 7620.0 | -3759.2 |
| ***C. Cubic polynomial (day, day², day³)*** | | | | |
|  | 2 | 8012.1 | 8034.5 | -3996.1 |
|  | 3 | 7776.6 | 7810.1 | -3873.3 |
|  | 4 | 7650.2 | 7694.8 | -3805.1 |
|  | 5 | 7598.4 | 7654.3 | -3774.2 |
|  | 6 | 7556.0 | 7623.0 | -3748.0 |

AIC, Akaike Information Criterion; BIC, Bayesian Information Criterion. Lower AIC/BIC values indicate improved model fit. The selected final model (quadratic polynomial, k=5 clusters; shaded) was chosen based on the combination of AIC/BIC improvement plateauing beyond 5 clusters and a priori clinical interpretability of the resulting phenotypes.

To assess the classification precision of the final 5-cluster solution, we calculated the average posterior probability of assignment (APPA)^6^ for each cluster, defined as the mean posterior probability of group membership among individuals whose hard classification (based on maximum posterior probability) fell in that cluster. APPA values ranged from 0.93 to 0.99 across the five phenotypes (c1: 0.94; c2: 0.93; c3: 0.96; c4: 0.987; c5: 0.99), all substantially exceeding the conventional threshold of 0.7 recommended for adequate classification quality^6^, indicating high assignment certainty across all clusters, including the smaller c4 and c5 subgroups (**Expanded Methods Figure M2**).

**Expanded Methods Figure M2. Average Posterior Probability of Assignment (APPA) for c1-c5 longitudinal mSOFA cluster phenotypes.**

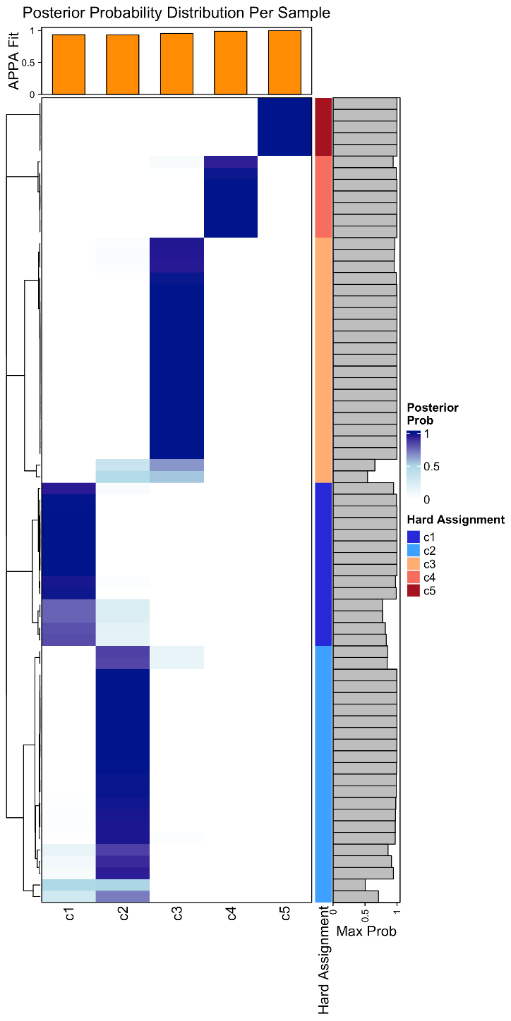

***4.2. Phenotype comparisons to demographic and clinical metrics***

Unless otherwise stated, all p-values were subjected to Benjamini-Hochberg (FDR) correction to control for type I error^7^. The five mSOFA phenotype clusters were statistically compared to demographic and outcome variables of interest. Age, Abbreviated Injury Scale/Injury Severity Score (AIS ISS) scores, lactate levels, and transfusion statistics were compared among mSOFA phenotype groups using analysis of variance (aov), Mann-Whitney U, χ^2^ or Fischer’s exact tests tests in R, as appropriate.

***4.3. Phenotype comparisons to O-link protein abundances***

Unless otherwise stated, all p-values were subjected to FDR correction to control for type I error. Protein abundances were modeled in R over the first 28 days of hospital admission using the kimma package (v1.6.3)^8^. Models included fixed effects for mSOFA group and time, the interaction between mSOFA group and time, and the random effect of subject ID.

***4.4. Statistical Citations***

1. Proust-Lima C, Philipps V, Diakite A, Liquet B (2025). *lcmm: Extended Mixed Models Using Latent Classes and Latent Processes*. R package version: 2.2.2, https://cran.r-project.org/package=lcmm.
2. R Core Team (2026). _R: A Language and Environment for Statistical Computing_. R Foundation for Statistical Computing, Vienna, Austria. doi:10.32614/R.manuals <https://doi.org/10.32614/R.manuals>. <https://www.R-project.org/>.
3. Akaike, H. (1974). A new look at the statistical model identification. *IEEE Transactions on Automatic Control*, 19(6), 716–723. https://doi.org/10.1109/TAC.1974.1100705.
4. Schwarz, Gideon E. (1978). "Estimating the dimension of a model." *Annals of Statistics*, 6 (2): 461–464.
5. Harville, D.A. (1974). Bayesian inference for variance components using only error contrasts. *Biometrika*, 61, 383--385. 10.2307/2334370.
6. Nagin DS, Odgers CL. Group-based trajectory modeling in clinical research. *Annu Rev Clin Psychol*. 2010;6:109-138.
7. Benjamini, Y., & Hochberg, Y. (1995). Controlling the false discovery rate: A practical and powerful approach to multiple testing. *Journal of the Royal Statistical Society: Series B (Methodological)*, 57(1), 289–300.
8. Dill-McFarland KA, Mitchell K, Batchu S, Segnitz RM, Benson B, Janczyk T, Cox MS, Mayanja-Kizza HA, Boom WH, Benchek P, Stein C, Hawn TR, Altman MC. 2023. kimma: Flexible linear mixed effects modeling with kinship for RNA-seq data. Bioinformatics. 39(5): btad279. doi: 10.1093/bioinformatics/btad279.

***5.1. Protein Function/Mechanism Annotation & Use of Artificial Intelligence Tools***

All proteins with significant differences in trajectory (FDR<0.1) across any of the analyses were annotated for function/mechanism by the investigators to assist and standardize phenotype biological interpretation. Initial Gene Ontology/PANTHER pathway analysis was performed but was deemed not to provide enough granularity and insight into this immunity-focused 92-analyte proteomic panel. Therefore, protein function/mechanism annotations were developed at the individual protein level by a combination of large language model platform literature review and human verification for accuracy and comprehensiveness. Literature identification and evidence synthesis were conducted using Consensus (consensus.app), an AI-powered academic search engine built on a GPT-4-based large language model fine-tuned on peer-reviewed scientific literature, which uses retrieval-augmented generation to return claim-level summaries with evidence-agreement scores from indexed papers. After literature synthesis and key reference manual review and vetting, annotations were finalized by the senior investigator (**Supplemental Table 1**).

Generative AI/AI-assisted tools were not used to collect, analyze or interpret any reported data. Claude AI (Model: Sonnet 5; Anthropic, PBC) was used for manuscript file management, document formatting, and suggested edits for brevity, readability and compliance with Journal manuscript submission requirements.
