## Supplementary material for "Multiple Organ Dysfunction Trajectory Phenotypes After Severe Trauma Are Associated With Distinct Immune Profiles and Outcomes": SDC3 – Supplemental Tables

**Supplemental Digital Content 3. Supplemental Tables**

**Table S1: Mechanism/Function Annotations for plasma immune markers significant (FDR < 0.1) in any phenotype analysis**

*Investigator-vetted mechanism/function annotations for the Olink Target 96 Inflammation panel proteins that reached FDR < 0.1 (n=67) in at least one of the five phenotype analyses performed (main effects across all clusters; overall time-interaction across all clusters; c1/c2/c3 vs c5 time-interaction; c1/c2/c3/c5 vs c4 time-interaction; c1/c2/c3 time-interaction). Proteins reported in SDC3 Tables S2 to S5 (n=46) derive from the four time-interaction analyses; the remaining 21 proteins reached FDR < 0.1 only in the main-effects analysis (phenotype difference in abundance independent of time), which is not reported in the manuscript, and are included for completeness. Proteins are listed alphabetically by protein/assay symbol. Up to 2 manually vetted supporting publication PMIDs are provided for each protein annotation. FDR, Benjamini-Hochberg false discovery rate; PMID, PubMed identifier.*

| **Protein/Assay Symbol** | **UniProt** | **Mechanism/Function Annotation** | **PMID(s)** |
| --- | --- | --- | --- |
| 4E-BP1 | Q13541 | Translational repressor downstream of mTORC1; phosphorylation of 4E-BP1 is constitutively elevated in regulatory T cells relative to naive T cells and is required for normal lymphocyte activation and proliferation during adaptive immune responses. | 23812589  35667842 |
| ADA | P00813 | Adenosine deaminase (ADA1/ADA2 isoenzymes) degrades extracellular adenosine, an immunosuppressive purine signal; by lowering adenosine levels ADA activity has a net pro-inflammatory effect, and the ADA2 isoenzyme specifically drives monocyte-to-macrophage differentiation and T-helper-cell proliferation. | 20453107  15926889 |
| ARTN | Q5T4W7 | Artemin, a GDNF-family neurotrophic ligand signaling through GFRα3/RET; human T cells, B cells, and monocytes express GDNF-family receptor components, and family ligands reduce TNF production by activated immune cells, indicating a neuroimmune modulatory role. | 16081799  33627484 |
| CASP-8 | Q14790 | Most upstream initiator protease of the extrinsic apoptosis cascade, recruited to death receptors (Fas, TNFR1, TRAIL-R) via FADD; beyond executing apoptosis, catalytically active caspase-8 also suppresses RIPK3/MLKL-dependent necroptosis and independently drives inflammatory cytokine production downstream of TLR and bacterial-infection signaling in innate immune cells. | 27737018  32531842 |
| CCL3 | P10147 | CC chemokine (MIP-1α) that recruits monocytes, macrophages, and T cells to sites of tissue injury via CCR1/CCR5 signaling, a core mediator of the early innate inflammatory response and, together with CCL4, preferentially engaged in Th1-type responses. | 33076281  15831559 |
| CCL4 | P13236 | MIP-1β, a CCR5 ligand that recruits CD8+ cytotoxic T lymphocytes to inflamed or tumor tissue, functionally distinct from CCR6-ligand CCL20 which preferentially recruits regulatory T cells. | 15831559  28553619 |
| CCL11 | P51671 | Eotaxin-1, a CCR3 ligand and potent eosinophil (and basophil/dendritic cell) chemoattractant; induced by TNF, IL-1α, and IFN-γ, CCL11 is central to allergic/type-2 tissue recruitment and has additional roles in fibroblast migration and cellular senescence. | 33846499 40429807 |
| CCL19 | Q99731 | CCR7 ligand and homeostatic chemokine that directs dendritic cell and naive/central-memory T cell trafficking to secondary lymphoid tissue. | 12077273  18379575 |
| CCL20 | P78556 | CCR6 ligand that recruits Th17 cells and immature dendritic cells to mucosal and inflamed tissue, linking innate epithelial signaling to adaptive Th17 responses. | 33076281  42284975 |
| CCL23 | P55773 | CCL23 (MPIF-1/CKβ8) is a CC chemokine that signals primarily through CCR1 to recruit monocytes, dendritic cells, and resting T lymphocytes to inflammatory sites. | 28553619  17371990 |
| CCL25 | O15444 | CCR9 ligand that mediates gut-homing chemotaxis of T lymphocytes, directing lymphocyte trafficking to intestinal mucosal tissue. | 33076281  42118152 |
| CCL28 | Q9NRJ3 | Mucosal-associated CC chemokine involved in recruitment of IgA-secreting plasma cells and other leukocytes to epithelial/mucosal surfaces. | 33076281  12538707 |
| CD5 | P06127 | Immunoregulatory receptor on T cells and B-1a cells that attenuates TCR/BCR signaling, protects autoreactive T cells from activation-induced cell death, and promotes peripheral induction of regulatory T cells. Soluble CD5 is hypothesized to function as a decoy receptor that blocks the interaction between membrane-bound CD5 and its endogenous ligands, thereby attenuating CD5-dependent survival and regulatory signals. | 30676652  24454761 |
| CD8A | P01732 | CD8 alpha chain, the T-cell receptor coreceptor for MHC class I:peptide complexes on cytotoxic T lymphocytes; strengthens TCR-antigen recognition by recruiting the Src kinase LCK, driving CD8+ T cell activation and cytotoxic effector function, and is used broadly as a marker/correlate of CD8+ T cell infiltration and activity. Circulating soluble CD8A correlates with the abundance of circulating cytotoxic and effector CD8+ T cells. | 39253084  41760714 |
| CD40 | P25942 | TNF-receptor-superfamily costimulatory receptor on antigen-presenting cells; CD40 signaling licenses dendritic cells, following cognate CD4+ T cell help, to prime effective CD8+ T cell (cytotoxic) responses. Plasma circulating CD40 functions as a soluble antagonist that dampens CD40L-driven humoral and inflammatory responses, and as a biomarker whose elevated levels reflect immune activation across autoimmune, malignant, and renal disease contexts. | 15475958  33202988 |
| CDCP1 | Q9H5V8 | CDCP1 (CD318), a transmembrane glycoprotein activating Src-family kinase signaling, is a documented ligand for the T-cell receptor CD6, alongside CD166 (ALCAM); this CD6/CDCP1 interaction is implicated in modulating T-cell adhesion and activation, though most detailed mechanistic data derive from cancer/epithelial contexts rather than direct lymphocyte-intrinsic studies. Treat immune-specific claims as exploratory. | 39996744  32983115 |
| CSF-1 | P09603 | Macrophage colony-stimulating factor, signaling through CSF1R, is the principal differentiation and survival factor for monocytes, macrophages, and osteoclasts and shapes tissue-resident macrophage phenotype. | 22186992 |
| CST5 | P28325 | Cystatin D, a cysteine protease inhibitor; direct immune-mechanistic literature is limited, but CST5 has been identified (along with 4E-BP1 and ST1A1) among the proteins most significantly associated with 28-day mortality after ST-elevation myocardial infarction in a study using the Olink 96 Inflammation panel. This annotation should be considered exploratory pending more specific mechanistic data. | 36209229 |
| CX3CL1 | P78423 | Fractalkine, the sole CX3C-family chemokine, exists in membrane-bound (adhesive) and soluble (chemotactic) forms and mediates monocyte, NK, and T cell adhesion and recruitment to activated endothelium. | 39125578  38731899 |
| CXCL1 | P09341 | ELR+ CXCR2 ligand and potent neutrophil chemoattractant, induced by IL-1β/TNF-α/NF-κB signaling, with a specifically documented role in acute kidney injury pathophysiology in addition to general inflammatory recruitment. Uncontrolled or sustained CXCL1-driven neutrophil recruitment causes collateral tissue damage across multiple organ systems. | 41898553  27625115 |
| CXCL5 | P42830 | ELR+ CXC chemokine and CXCR2 ligand that drives neutrophil trafficking to injured or infected tissue; rapidly induced by IL-1β/TNF-α in endothelial and epithelial cells and mechanistically implicated in epithelial barrier defense; a plasma proteomics GWAS causally implicated CXCL5 in mucosal barrier disease (ulcerative colitis). | 17878384  37563310 |
| CXCL6 | P80162 | GCP-2, an ELR+ CXC chemokine that, unusually among the CXCR2 ligands, binds both CXCR1 and CXCR2, giving it a broad neutrophil-activating profile. Unlike newly-synthesized CXCL8, CXCL6 is stored preformed in neutrophil granules and released immediately on activation, and it also retains direct antimicrobial activity. | 25993443  12628493 |
| CXCL9 | Q07325 | IFN-γ-induced CXCR3 ligand (Mig) that, together with CXCL10 and CXCL11, recruits effector Th1/CD8+ T cells to sites of inflammation. It is among the CXCR3-ligand amplifiers of type 1 immunity by recruiting effector T cells, driving Th1 polarization, and serving as a systemic biomarker for IFN-γ activity and immunotherapy response. | 26657511  21221121 |
| CXCL10 | P02778 | IFN-γ-induced CXCR3 ligand (IP-10) that recruits activated Th1 cells and cytotoxic T lymphocytes; classified as a "driver chemokine" of inflammatory recruitment. | 9064356  26657511 |
| FGF-5 | Q8NF90 | Paracrine FGFR1 ligand; organ-specific and immune-specific mechanistic literature is limited (best-characterized role is regulation of the hair follicle growth cycle in animal models). General FGF-family class effects (fibroblast proliferation, tissue remodeling) likely apply. This annotation should be treated as exploratory. | 27036966  34831463 |
| FGF-21 | Q9NSA1 | Fibroblast growth factor 21 (FGF21) is a stress-inducible hepatokine that circulates to distant target tissues (brain, adipose tissue, and skeletal muscle) to coordinate systemic responses to nutritional stress. It also elevated in metabolic disease (obesity, T2DM) and acts as an insulin sensitizer. | 35301051  25008183 |
| FGF-23 | Q9GZV9 | A phosphaturic hormone whose circulating levels rise with declining renal function; FGF23 also directly stimulates hepatic and systemic inflammatory cytokine production via FGFR4, and its production is itself upregulated by TNF and IL-1β, creating a feed-forward inflammation-FGF23 loop. | 31301888  36624273 |
| Flt3L | P49771 | Hematopoietic growth factor that expands early progenitor populations and drives generation of conventional and plasmacytoid dendritic cells and NK cells; essential for normal dendritic cell and NK cell numbers in vivo. | 10910900  38701783 |
| GDNF | P39905 | Glial-cell-line-derived neurotrophic factor; human T cells, B cells, and monocytes express the RET/GFRα receptor components for this ligand family, and GDNF-family signaling reduces TNF production by activated immune cells, indicating a direct neuroimmune modulatory function beyond its classical neurotrophic role. | 16081799  11988777 |
| HGF | P14210 | Hepatocyte growth factor promotes differentiation of monocytes into regulatory, IL-10-high/IL-12-low accessory cells that induce FoxP3+ regulatory T cells, and directly suppresses macrophage IL-6 production while enhancing IL-10, indicating a net immunoregulatory/anti-inflammatory function. | 16527888  21072211 |
| IL-1 alpha | P01583 | Founding member of the IL-1 cytokine family and a prototypical dual-function alarmin; constitutively expressed and chromatin-bound intracellularly, IL-1α is released from necrotic (but not apoptotic) cells to bind IL-1R1 on neighboring cells and trigger sterile inflammation, making it a direct molecular signal of tissue injury and cell death rather than a pathogen-triggered cytokine. | 30995499  29247995 |
| IL-8 | P10145 | CXCL8/IL-8 is the most potent human neutrophil chemoattractant, signaling through CXCR1/CXCR2 to drive neutrophil recruitment and activation at sites of infection or injury. | 36725964  41736410 |
| IL-10RA | Q13651 | Ligand-binding α-chain of the IL-10 receptor, paired with IL-10RB; IL-10RA engagement by IL-10 activates JAK1/STAT3 signaling that inhibits proinflammatory and co-stimulatory function in innate and adaptive immune cells, and loss-of-function mutations in this receptor chain cause very-early-onset inflammatory bowel disease. In severe trauma, circulating IL-10 peaks within minutes at the injury scene, preceding the rise of pro-inflammatory cytokines and correlating with HLA-DR suppression. | 33737461  26912315 |
| IL-10RB | Q08334 | Shared β-chain receptor subunit used by the IL-10, IL-22, IL-26, and type III interferon (IL-28/29) signaling complexes; required alongside the respective α-chain for JAK/STAT-mediated signal transduction. | 29016674  32229722 |
| IL-15RA | Q13261 | High-affinity receptor subunit that chaperones and trans-presents IL-15 to neighboring NK cells and memory CD8+ T cells; this trans-presentation mechanism is the primary driver of NK and memory CD8+ T cell homeostasis in vivo. The soluble receptor retains high affinity for IL-15 and behaves as a potent, specific inhibitor of IL-15 binding to the membrane receptor and of IL-15-induced cell proliferation. | 19913445  15265897 |
| IL-17A | Q16552 | Canonical Th17-lineage effector cytokine that promotes neutrophil recruitment and antimicrobial peptide production for protective immunity against bacterial and fungal pathogens, but drives immunopathology when dysregulated in chronic inflammation and autoimmunity (e.g., psoriasis). | 35790881  31681282 |
| IL-17C | Q9P0M4 | Epithelial-cell-derived member of the IL-17 family that acts in an autocrine loop via a distinct receptor (IL-17RA/IL-17RE) expressed on epithelium rather than leukocytes, driving rapid innate antimicrobial and inflammatory gene expression at barrier surfaces. mechanistically distinct from T-cell-derived IL-17A. Plasma circulating IL-17C functions as an early epithelial alarmin that reinforces innate barrier defense and amplifies adaptive Th17 responses. | 21993848  23221046 |
| IL-18 | Q14116 | IL-1-family cytokine that induces IFN-γ production from Th1 cells and NK cells (especially with IL-12), with additional pleiotropic actions on mast cells, basophils, and CD8+ T cells. In severe trauma, circulating IL-18 rises rapidly due to constitutive precursor availability in injured tissues, and its persistent elevation serves as a robust biomarker for ARDS, sepsis, and multi-organ dysfunction | 38081945  19054512 |
| IL-18R1 | Q13478 | Ligand-binding chain of the IL-18 receptor complex; co-expression with the IL-18RAP accessory chain is required for MyD88-dependent activation of NF-κB and MAPK signaling downstream of IL-18. | 29247988  39769266 |
| IL-20RA | Q9UHF4 | Receptor α-chain (with IL-20RB) for the IL-20 subfamily (IL-19, IL-20, IL-24); expressed on immune and epithelial cells, IL-20R cytokine signaling is generally associated with chronic inflammation and autoimmune disease, though individual family members (e.g., IL-19) can be anti-inflammatory, giving this receptor complex context-dependent immunoregulatory function. | 30260500  12351624 |
| IL-22RA1 | Q8N6P7 | Receptor α-subunit shared by IL-20, IL-22, and IL-24 signaling complexes, expressed selectively on epithelial cells. IL-22RA1 mediates epithelial barrier defense and tissue repair via STAT3, and lower circulating levels of its ligand IL-22 after trauma have been associated with greater injury severity and respiratory failure requiring intubation. | 30319661  40857975 |
| IL-24 | Q13007 | Member of the IL-20 cytokine subfamily signaling through IL-20R/IL-22R receptor complexes; produced by monocytes in response to TLR ligands and immune complexes. In severely injured trauma patients, IL-24 belongs to a subset of protective and reparative cytokines that are suppressed at high injury severity, a pattern associated with worse clinical outcomes. | 30319661  33748428 |
| IL-33 | O95760 | Nuclear-associated IL-1-family alarmin constitutively expressed in endothelial and epithelial cells; released upon cell injury or necrosis, IL-33 signals through its receptor ST2 (IL-1RL1) to activate mast cells, ILC2s, Th2 cells, and a highly suppressive ST2+ regulatory T cell subset, giving it dual pro-inflammatory (type 2 immunity, tissue remodeling/fibrosis) and tissue-repair/immunoregulatory functions depending on chronicity and cellular context. | 29247993  38082335 |
| IL6 | P05231 | Pleiotropic cytokine signaling via classic (membrane IL-6R) and trans- (soluble IL-6R) pathways; central regulator of the Th17/regulatory T cell balance and a key driver of the acute-phase and systemic inflammatory response. IL-6 drives the acute-phase response and predicts organ failure after severe trauma, but excessive elevation triggers immunosuppression and mortality. | 10722030  20583029 |
| IL10 | P22301 | Principal anti-inflammatory cytokine that inhibits proinflammatory and co-stimulatory function of innate and adaptive immune cells; essential for maintaining mucosal immune homeostasis. IL-10 suppresses pro-inflammatory immunity to prevent tissue damage, but its early surge after severe trauma drives immunosuppression and predicts sepsis and mortality. | 33717127  8614042 |
| LAP TGF-beta-1 | P01137 | Latency-associated peptide of TGF-β1 regulates the bioavailability of active TGF-β1, a cytokine central to regulatory T cell induction and restraint of effector T cell expansion; TGF-β signaling works alongside PD-L1 to limit stem-like CD8+ T cell expansion in inflamed tissue. Elevated circulating TGF-β1 after trauma drives immunosuppression, and antibody blockade restores lymphocyte proliferation and IL-2 production. | 37543621  8656475 |
| LIF | P15018 | IL-6-family pleiotropic cytokine with immunosuppressive activity in inflamed/tumor tissue; elevated circulating LIF is associated with impaired CD8+ T cell infiltration and poorer immune-mediated outcomes. | 33259884  34416362 |
| LIF-R | P42702 | Receptor for LIF that, in complex with gp130, activates JAK/STAT3 signaling; regulates a broad array of context-dependent tumor- and tissue-related processes including proliferation and cell survival, and forms the type I receptor complex shared with the related cytokine OSM. | 39279155 |
| MCP-1 | P13500 | CCL2 is the principal monocyte-chemoattractant chemokine, produced by endothelium in response to IL-1β/TNF-α, signaling through CCR2 to drive monocyte, NK, and CD4+/CD8+ T cell infiltration into injured or infected tissue and promoting Th2-biased responses. | 37208442  37720514 |
| MCP-3 | P80098 | CCL7 is a monocyte and eosinophil chemoattractant that synergizes with CXCL8 to enhance neutrophil chemotaxis during inflammation. Plasma MCP-3 rises significantly within 6 to 24 hours post-injury in mice, consistent with a role as an acute-phase reactant that signals tissue damage and initiates immune cell infiltration. | 15075362  34833127 |
| MMP-1 | P03956 | Interstitial collagenase that degrades fibrillar collagens (types I, II, III) as part of extracellular matrix turnover during tissue remodeling and repair; regulation of MMP family activity, latency, and tissue inhibitors (TIMPs) is reviewed extensively in the matrix biology literature. | 16405877 |
| MMP-10 | P09238 | Zinc-dependent matrix metalloproteinase involved in tissue remodeling and apoptosis regulation; in acute kidney injury models MMP-10 is protective, augmenting EGFR-AKT/ERK signaling to promote tubular cell survival and regeneration. In severe systemic inflammation and trauma, circulating MMP-10 rises acutely and correlates with disease severity. | 33436543  19799791 |
| NRTN | Q99748 | Neurturin, a GDNF-family ligand signaling via GFRα2/RET; produced by activated human T cells, B cells, and monocytes, and reduces TNF protein production by activated PBMCs without altering transcription, suggesting a post-transcriptional immunomodulatory role. | 16081799 32252363 |
| NT-3 | P20783 | Neurotrophin-3, a target-derived neurotrophic factor best characterized for sensory/proprioceptive neuron survival and development; NT-3 signals primarily through the TrkC receptor, activating downstream MAPK, PI3K/Akt, and Erk5 pathways to modulate both neuronal survival and suppresses monocyte and T-cell activation while also driving macrophage-mediated tissue repair. | 18657279  34688600 |
| OPG | O00300 | Osteoprotegerin (TNFRSF11B) is a soluble TNF-receptor-superfamily decoy receptor for RANKL that regulates dendritic cell and osteoclast survival and is released by endothelium in response to inflammatory stimuli, linking it to vascular inflammation and injury. | 30736365  23827649 |
| PD-L1 | Q9NZQ7 | Immune checkpoint ligand that engages PD-1 on activated T cells to counteract T cell receptor activating signals, promoting immune tolerance and, in chronic inflammation or malignancy, immune evasion. Soluble programmed death-ligand 1 (sPD-L1) is a shed, circulating form of the membrane-bound checkpoint ligand that retains its ability to bind PD-1 on immune cells, thereby inducing immune exhaustion and suppressing T-cell effector functions. | 34575296  40208036 |
| SIRT2 | Q8IXJ6 | NAD+-dependent deacetylase, the principal cytoplasmic sirtuin in myeloid cells; SIRT2 restrains inflammation by deacetylating and inhibiting the NF-κB p65 subunit. In the innate immune system, SIRT2 is the most highly expressed sirtuin in myeloid cells and exerts crucial regulatory effects on macrophage activation and immune homeostasis. | 32768387  25072851 |
| SLAMF1 | Q13291 | CD150, a self-ligand receptor of the SLAM family expressed broadly on hematopoietic cells; bridges innate and adaptive immunity by supporting T cell costimulation/proliferation and, together with the SAP adaptor, mediating phagosome maturation and bacterial sensing in phagocytes. | 40701970  32983115 |
| ST1A1 | P50225 | Sulfotransferase 1A1, a phase II conjugating enzyme for catecholamines and other substrates; direct immune-mechanistic literature is limited, but ST1A1 (along with CST5 and 4E-BP1) was among the proteins most significantly associated with 28-day mortality after ST-elevation myocardial infarction in a study using the Olink 96 Inflammation panel. This annotation should be treated as exploratory pending more specific mechanistic data. | 36209229 |
| STAMBP | O95630 | STAM-binding protein, a deubiquitinase that constrains NLRP3 (and NALP7) inflammasome activation; loss of STAMBP increases IL-1β release and broadly exaggerates cytokine/chemokine production in response to TLR/LPS stimulation, indicating a physiologic braking function on innate inflammasome-driven inflammation. | 33253913  28492230 |
| TGF-alpha | P01135 | EGFR ligand primarily characterized for epithelial proliferation and wound repair; direct immune-specific mechanistic literature is limited. Treat this annotation as exploratory and unconfirmed pending protein-specific immune data. | 40722787 |
| TNF | P01375 | Master pro-inflammatory cytokine (TNF-alpha) produced principally by activated macrophages; drives the systemic inflammatory response and, through TNFR2 signaling, also amplifies regulatory T cell expansion and suppressive activity, illustrating its dual pro-inflammatory/immunoregulatory roles. | 39745933  21491419 |
| TNFRSF9 | Q07011 | 4-1BB/CD137, a TNF-receptor-superfamily costimulatory receptor induced on activated T and NK cells; enhances CD8+ T cell proliferation, survival, and effector cytokine production following TCR engagement. | 39745933  9206996 |
| TSLP | Q969D9 | Epithelial-cell-derived cytokine that activates dendritic cells to drive type 2 (Th2) immune responses, linking barrier epithelial signaling to innate and adaptive immune activation; also has emerging roles in infectious and inflammatory disease beyond classic allergy. | 35650271  20300138 |
| TWEAK | O43508 | TNFSF12, a TNF-superfamily cytokine signaling through its sole receptor Fn14 (TNFRSF12A); TWEAK/Fn14 activates canonical and non-canonical NF-κB signaling to drive inflammatory cytokine/chemokine production, cell proliferation, and angiogenesis, functioning as a physiologic mediator of tissue repair after acute injury but promoting pathological inflammation and fibrosis when chronically dysregulated, including in kidney injury. | 35650271  16325585 |
| uPA | P00749 | Circulating urokinase-type plasminogen activator (uPA) is a serine protease that converts plasminogen to plasmin, linking fibrinolysis to extracellular matrix degradation and neutrophil trafficking during tissue injury and repair. | 35580335  21969013 |
| VEGFA | P15692 | Angiogenic growth factor with biphasic expression during acute kidney injury, an early peak supporting peritubular capillary maintenance and a later peak associated with the transition to chronic injury and fibrotic repair. Beyond its role in angiogenesis and renal microvascular protection, VEGF-A acts as a potent immunosuppressive factor by directly modulating myeloid cells, dendritic cells, and T cell function through VEGFR2 signaling. | 37055531  35466428 |

**Table S2. Phenotypes c1/c2/c3/c4/c5 plasma immune proteomic analysis**

*Plasma immune marker proteins (n=40) significant in the mixed-effects time-interaction analysis across the five mSOFA trajectory phenotypes (c1-c5), selected for FDR < 0.1, ranked from most to least significant. FDR, Benjamini-Hochberg false discovery rate.*

| **Protein/Assay Symbol** | **FDR** | **Protein/Assay Symbol** | **FDR** |
| --- | --- | --- | --- |
| CD5 | 1.05E-8 | IL-24 | 0.0211 |
| IL-15RA | 1.55E-6 | TWEAK | 0.0211 |
| FGF-5 | 5.31E-5 | uPA | 0.0257 |
| CD8A | 5.21E-4 | MMP-10 | 0.0324 |
| SIRT2 | 5.52E-4 | CXCL5 | 0.0336 |
| IL-20RA | 9.04E-4 | CST5 | 0.0392 |
| ARTN | 0.0013 | 4E-BP1 | 0.0394 |
| CX3CL1 | 0.0017 | CXCL9 | 0.0403 |
| ADA | 0.0023 | IL-18 | 0.057 |
| CCL28 | 0.0023 | IL-10RB | 0.0641 |
| VEGFA | 0.0031 | IL-22RA1 | 0.0684 |
| STAMBP | 0.0047 | CXCL6 | 0.0734 |
| FGF-23 | 0.0101 | LAP TGF-beta-1 | 0.0797 |
| HGF | 0.0101 | LIF-R | 0.0797 |
| NT-3 | 0.0111 | MMP-1 | 0.0797 |
| ST1A1 | 0.0119 | MCP-1 | 0.0824 |
| CCL11 | 0.012 | CASP-8 | 0.0829 |
| TNFRSF9 | 0.0133 | IL-1 alpha | 0.0829 |
| IL-17C | 0.0187 | CCL25 | 0.0842 |
| OPG | 0.0187 | CSF-1 | 0.088 |

**Table S3. Phenotypes c5 vs c1/c2/c3 plasma immune proteomic analysis**

*Plasma immune marker proteins (n=3) significant in the mixed-effects time-interaction analysis specific to the c1/c2/c3 vs c5 mSOFA trajectory phenotype contrast, FDR < 0.1, ranked from most to least significant, with mechanism/function annotations and supporting PMIDs. Model coefficient is for c5 with c1/c2/c3 as reference (thus negative when c5 slope is lower). FDR, Benjamini-Hochberg false discovery rate.*

| **Protein/Assay Symbol** | **Model Coefficient** | **FDR** | **Mechanism/Function Annotation** | **PMID(s)** |
| --- | --- | --- | --- | --- |
| IL-17C | -0.098 | 0.0734 | Epithelial-cell-derived member of the IL-17 family that acts in an autocrine loop via a distinct receptor (IL-17RA/IL-17RE) expressed on epithelium rather than leukocytes, driving rapid innate antimicrobial and inflammatory gene expression at barrier surfaces. mechanistically distinct from T-cell-derived IL-17A. Plasma circulating IL-17C functions as an early epithelial alarmin that reinforces innate barrier defense and amplifies adaptive Th17 responses. | 21993848  23221046 |
| CXCL5 | -0.089 | 0.0866 | ELR+ CXC chemokine and CXCR2 ligand that drives neutrophil trafficking to injured or infected tissue; rapidly induced by IL-1β/TNF-α in endothelial and epithelial cells and mechanistically implicated in epithelial barrier defense; a plasma proteomics GWAS causally implicated CXCL5 in mucosal barrier disease (ulcerative colitis). | 17878384  37563310 |
| CXCL6 | -0.058 | 0.0866 | GCP-2, an ELR+ CXC chemokine that, unusually among the CXCR2 ligands, binds both CXCR1 and CXCR2, giving it a broad neutrophil-activating profile. Unlike newly-synthesized CXCL8, CXCL6 is stored preformed in neutrophil granules and released immediately on activation, and it also retains direct antimicrobial activity. | 25993443  12628493 |

**Table S4. Phenotype c1/c2/c3 plasma immune proteomic analysis**

*Plasma immune marker proteins (n=13) significant in the mixed-effects time-interaction analysis among the low-severity c1/c2/c3 mSOFA trajectory phenotypes, FDR < 0.1, ranked from most to least significant, with mechanism/function annotations and supporting PMIDs. Model coefficient is for c3 with c1/c2 as reference (thus positive when c3 slope is greater). FDR, Benjamini-Hochberg false discovery rate.*

| **Protein/Assay Symbol** | **Model Coefficient** | **FDR** | **Mechanism/Function Annotation** | **PMID(s)** |
| --- | --- | --- | --- | --- |
| ADA | -0.0601 | 0.0121 | Adenosine deaminase (ADA1/ADA2 isoenzymes) degrades extracellular adenosine, an immunosuppressive purine signal; by lowering adenosine levels ADA activity has a net pro-inflammatory effect, and the ADA2 isoenzyme specifically drives monocyte-to-macrophage differentiation and T-helper-cell proliferation. | 20453107  15926889 |
| CCL11 | 0.0328 | 0.0121 | Eotaxin-1, a CCR3 ligand and potent eosinophil (and basophil/dendritic cell) chemoattractant; induced by TNF, IL-1α, and IFN-γ, CCL11 is central to allergic/type-2 tissue recruitment and has additional roles in fibroblast migration and cellular senescence. | 33846499 40429807 |
| SIRT2 | -0.1193 | 0.0121 | NAD+-dependent deacetylase, the principal cytoplasmic sirtuin in myeloid cells; SIRT2 restrains inflammation by deacetylating and inhibiting the NF-κB p65 subunit. In the innate immune system, SIRT2 is the most highly expressed sirtuin in myeloid cells and exerts crucial regulatory effects on macrophage activation and immune homeostasis. | 32768387  25072851 |
| STAMBP | -0.0982 | 0.0121 | STAM-binding protein, a deubiquitinase that constrains NLRP3 (and NALP7) inflammasome activation; loss of STAMBP increases IL-1β release and broadly exaggerates cytokine/chemokine production in response to TLR/LPS stimulation, indicating a physiologic braking function on innate inflammasome-driven inflammation. | 33253913  28492230 |
| TWEAK | 0.0427 | 0.0180 | TNFSF12, a TNF-superfamily cytokine signaling through its sole receptor Fn14 (TNFRSF12A); TWEAK/Fn14 activates canonical and non-canonical NF-κB signaling to drive inflammatory cytokine/chemokine production, cell proliferation, and angiogenesis, functioning as a physiologic mediator of tissue repair after acute injury but promoting pathological inflammation and fibrosis when chronically dysregulated, including in kidney injury. | 35650271  16325585 |
| CST5 | 0.0516 | 0.0183 | Cystatin D, a cysteine protease inhibitor; direct immune-mechanistic literature is limited, but CST5 has been identified (along with 4E-BP1 and ST1A1) among the proteins most significantly associated with 28-day mortality after ST-elevation myocardial infarction in a study using the Olink 96 Inflammation panel. This annotation should be considered exploratory pending more specific mechanistic data. | 36209229 |
| HGF | 0.0715 | 0.0326 | Hepatocyte growth factor promotes differentiation of monocytes into regulatory, IL-10-high/IL-12-low accessory cells that induce FoxP3+ regulatory T cells, and directly suppresses macrophage IL-6 production while enhancing IL-10, indicating a net immunoregulatory/anti-inflammatory function. | 16527888  21072211 |
| IL-24 | 0.0616 | 0.0326 | Member of the IL-20 cytokine subfamily signaling through IL-20R/IL-22R receptor complexes; produced by monocytes in response to TLR ligands and immune complexes. In severely injured trauma patients, IL-24 belongs to a subset of protective and reparative cytokines that are suppressed at high injury severity, a pattern associated with worse clinical outcomes. | 30319661  33748428 |
| ST1A1 | -0.0433 | 0.0326 | Sulfotransferase 1A1, a phase II conjugating enzyme for catecholamines and other substrates; direct immune-mechanistic literature is limited, but ST1A1 (along with CST5 and 4E-BP1) was among the proteins most significantly associated with 28-day mortality after ST-elevation myocardial infarction in a study using the Olink 96 Inflammation panel. This annotation should be treated as exploratory pending more specific mechanistic data. | 36209229 |
| ARTN | 0.0309 | 0.0431 | Artemin, a GDNF-family neurotrophic ligand signaling through GFRα3/RET; human T cells, B cells, and monocytes express GDNF-family receptor components, and family ligands reduce TNF production by activated immune cells, indicating a neuroimmune modulatory role. | 16081799  33627484 |
| 4E-BP1 | -0.1042 | 0.0743 | Translational repressor downstream of mTORC1; phosphorylation of 4E-BP1 is constitutively elevated in regulatory T cells relative to naive T cells and is required for normal lymphocyte activation and proliferation during adaptive immune responses. | 23812589  35667842 |
| CASP-8 | -0.0635 | 0.0784 | Most upstream initiator protease of the extrinsic apoptosis cascade, recruited to death receptors (Fas, TNFR1, TRAIL-R) via FADD; beyond executing apoptosis, catalytically active caspase-8 also suppresses RIPK3/MLKL-dependent necroptosis and independently drives inflammatory cytokine production downstream of TLR and bacterial-infection signaling in innate immune cells. | 27737018  32531842 |
| CXCL9 | 0.0440 | 0.0791 | IFN-γ-induced CXCR3 ligand (Mig) that, together with CXCL10 and CXCL11, recruits effector Th1/CD8+ T cells to sites of inflammation. It is among the CXCR3-ligand amplifiers of type 1 immunity by recruiting effector T cells, driving Th1 polarization, and serving as a systemic biomarker for IFN-γ activity and immunotherapy response. | 26657511  21221121 |

**Table S5. Phenotypes c1/c2/c3/c5 vs c4 plasma immune proteomic analysis**

*Plasma immune marker proteins (n=29) significant in the mixed-effects time-interaction analysis specific to the c1/c2/c3/c5 vs c4 mSOFA trajectory phenotype contrast, FDR < 0.1, ranked from most to least significant, with mechanism/function annotations and supporting PMIDs. Model coefficient is for c4 with c1/c2/c3/c5 as reference (thus positive when c4 slope is greater). FDR, Benjamini-Hochberg false discovery rate.*

| **Protein/Assay Symbol** | **Model Coefficient** | **FDR** | **Mechanism/Function Annotation** | **PMID(s)** |
| --- | --- | --- | --- | --- |
| CD5 | 0.0696 | 4.47E-10 | Immunoregulatory receptor on T cells and B-1a cells that attenuates TCR/BCR signaling, protects autoreactive T cells from activation-induced cell death, and promotes peripheral induction of regulatory T cells. Soluble CD5 is hypothesized to function as a decoy receptor that blocks the interaction between membrane-bound CD5 and its endogenous ligands, thereby attenuating CD5-dependent survival and regulatory signals. | 30676652  24454761 |
| IL-15RA | 0.0673 | 7.25E-7 | High-affinity receptor subunit that chaperones and trans-presents IL-15 to neighboring NK cells and memory CD8+ T cells; this trans-presentation mechanism is the primary driver of NK and memory CD8+ T cell homeostasis in vivo. The soluble receptor retains high affinity for IL-15 and behaves as a potent, specific inhibitor of IL-15 binding to the membrane receptor and of IL-15-induced cell proliferation. | 19913445  15265897 |
| FGF-5 | 0.0442 | 1.62E-5 | Paracrine FGFR1 ligand; organ-specific and immune-specific mechanistic literature is limited (best-characterized role is regulation of the hair follicle growth cycle in animal models). General FGF-family class effects (fibroblast proliferation, tissue remodeling) likely apply. This annotation should be treated as exploratory. | 27036966  34831463 |
| CD8A | 0.0770 | 1.47E-4 | CD8 alpha chain, the T-cell receptor coreceptor for MHC class I:peptide complexes on cytotoxic T lymphocytes; strengthens TCR-antigen recognition by recruiting the Src kinase LCK, driving CD8+ T cell activation and cytotoxic effector function, and is used broadly as a marker/correlate of CD8+ T cell infiltration and activity. Circulating soluble CD8A correlates with the abundance of circulating cytotoxic and effector CD8+ T cells. | 39253084  41760714 |
| VEGFA | 0.0654 | 8.99E-4 | Angiogenic growth factor with biphasic expression during acute kidney injury, an early peak supporting peritubular capillary maintenance and a later peak associated with the transition to chronic injury and fibrotic repair. Beyond its role in angiogenesis and renal microvascular protection, VEGF-A acts as a potent immunosuppressive factor by directly modulating myeloid cells, dendritic cells, and T cell function through VEGFR2 signaling. | 37055531  35466428 |
| CX3CL1 | 0.0589 | 0.0015 | Fractalkine, the sole CX3C-family chemokine, exists in membrane-bound (adhesive) and soluble (chemotactic) forms and mediates monocyte, NK, and T cell adhesion and recruitment to activated endothelium. | 39125578  38731899 |
| IL-20RA | 0.0487 | 0.0019 | Receptor α-chain (with IL-20RB) for the IL-20 subfamily (IL-19, IL-20, IL-24); expressed on immune and epithelial cells, IL-20R cytokine signaling is generally associated with chronic inflammation and autoimmune disease, though individual family members (e.g., IL-19) can be anti-inflammatory, giving this receptor complex context-dependent immunoregulatory function. | 30260500  12351624 |
| TNFRSF9 | 0.0552 | 0.0044 | 4-1BB/CD137, a TNF-receptor-superfamily costimulatory receptor induced on activated T and NK cells; enhances CD8+ T cell proliferation, survival, and effector cytokine production following TCR engagement. | 39745933  9206996 |
| ARTN | 0.0661 | 0.0062 | Artemin, a GDNF-family neurotrophic ligand signaling through GFRα3/RET; human T cells, B cells, and monocytes express GDNF-family receptor components, and family ligands reduce TNF production by activated immune cells, indicating a neuroimmune modulatory role. | 16081799  33627484 |
| FGF-23 | 0.1671 | 0.0062 | A phosphaturic hormone whose circulating levels rise with declining renal function; FGF23 also directly stimulates hepatic and systemic inflammatory cytokine production via FGFR4, and its production is itself upregulated by TNF and IL-1β, creating a feed-forward inflammation-FGF23 loop. | 31301888  36624273 |
| OPG | 0.0581 | 0.0065 | Osteoprotegerin (TNFRSF11B) is a soluble TNF-receptor-superfamily decoy receptor for RANKL that regulates dendritic cell and osteoclast survival and is released by endothelium in response to inflammatory stimuli, linking it to vascular inflammation and injury. | 30736365  23827649 |
| CCL28 | 0.1112 | 0.0107 | Mucosal-associated CC chemokine involved in recruitment of IgA-secreting plasma cells and other leukocytes to epithelial/mucosal surfaces. | 33076281  12538707 |
| LIF-R | 0.0298 | 0.0273 | Receptor for LIF that, in complex with gp130, activates JAK/STAT3 signaling; regulates a broad array of context-dependent tumor- and tissue-related processes including proliferation and cell survival, and forms the type I receptor complex shared with the related cytokine OSM. | 39279155 |
| uPA | 0.0365 | 0.0277 | Circulating urokinase-type plasminogen activator (uPA) is a serine protease that converts plasminogen to plasmin, linking fibrinolysis to extracellular matrix degradation and neutrophil trafficking during tissue injury and repair. | 35580335  21969013 |
| NT-3 | 0.0487 | 0.0400 | Neurotrophin-3, a target-derived neurotrophic factor best characterized for sensory/proprioceptive neuron survival and development; NT-3 signals primarily through the TrkC receptor, activating downstream MAPK, PI3K/Akt, and Erk5 pathways to modulate both neuronal survival and suppresses monocyte and T-cell activation while also driving macrophage-mediated tissue repair. | 18657279  34688600 |
| IL-10RB | 0.0281 | 0.0414 | Shared β-chain receptor subunit used by the IL-10, IL-22, IL-26, and type III interferon (IL-28/29) signaling complexes; required alongside the respective α-chain for JAK/STAT-mediated signal transduction. | 29016674  32229722 |
| PD-L1 | 0.0444 | 0.0455 | Immune checkpoint ligand that engages PD-1 on activated T cells to counteract T cell receptor activating signals, promoting immune tolerance and, in chronic inflammation or malignancy, immune evasion. Soluble programmed death-ligand 1 (sPD-L1) is a shed, circulating form of the membrane-bound checkpoint ligand that retains its ability to bind PD-1 on immune cells, thereby inducing immune exhaustion and suppressing T-cell effector functions. | 34575296  40208036 |
| IL-22RA1 | 0.0636 | 0.0489 | Receptor α-subunit shared by IL-20, IL-22, and IL-24 signaling complexes, expressed selectively on epithelial cells. IL-22RA1 mediates epithelial barrier defense and tissue repair via STAT3, and lower circulating levels of its ligand IL-22 after trauma have been associated with greater injury severity and respiratory failure requiring intubation. | 30319661  40857975 |
| CCL25 | 0.0409 | 0.0548 | CCR9 ligand that mediates gut-homing chemotaxis of T lymphocytes, directing lymphocyte trafficking to intestinal mucosal tissue. | 33076281  42118152 |
| MMP-10 | 0.0526 | 0.0548 | Zinc-dependent matrix metalloproteinase involved in tissue remodeling and apoptosis regulation; in acute kidney injury models MMP-10 is protective, augmenting EGFR-AKT/ERK signaling to promote tubular cell survival and regeneration. In severe systemic inflammation and trauma, circulating MMP-10 rises acutely and correlates with disease severity. | 33436543  19799791 |
| CD40 | 0.0453 | 0.0583 | TNF-receptor-superfamily costimulatory receptor on antigen-presenting cells; CD40 signaling licenses dendritic cells, following cognate CD4+ T cell help, to prime effective CD8+ T cell (cytotoxic) responses. Plasma circulating CD40 functions as a soluble antagonist that dampens CD40L-driven humoral and inflammatory responses, and as a biomarker whose elevated levels reflect immune activation across autoimmune, malignant, and renal disease contexts. | 15475958  33202988 |
| IL-10RA | 0.0382 | 0.0605 | Ligand-binding α-chain of the IL-10 receptor, paired with IL-10RB; IL-10RA engagement by IL-10 activates JAK1/STAT3 signaling that inhibits proinflammatory and co-stimulatory function in innate and adaptive immune cells, and loss-of-function mutations in this receptor chain cause very-early-onset inflammatory bowel disease. In severe trauma, circulating IL-10 peaks within minutes at the injury scene, preceding the rise of pro-inflammatory cytokines and correlating with HLA-DR suppression. | 33737461  26912315 |
| TGF-alpha | 0.0641 | 0.0653 | EGFR ligand primarily characterized for epithelial proliferation and wound repair; direct immune-specific mechanistic literature is limited. Treat this annotation as exploratory and unconfirmed pending protein-specific immune data. | 40722787 |
| MCP-1 | 0.0489 | 0.0747 | CCL2 is the principal monocyte-chemoattractant chemokine, produced by endothelium in response to IL-1β/TNF-α, signaling through CCR2 to drive monocyte, NK, and CD4+/CD8+ T cell infiltration into injured or infected tissue and promoting Th2-biased responses. | 37208442  37720514 |
| CSF-1 | 0.0167 | 0.0791 | Macrophage colony-stimulating factor, signaling through CSF1R, is the principal differentiation and survival factor for monocytes, macrophages, and osteoclasts and shapes tissue-resident macrophage phenotype. | 22186992 |
| SIRT2 | 0.0748 | 0.0810 | NAD+-dependent deacetylase, the principal cytoplasmic sirtuin in myeloid cells; SIRT2 restrains inflammation by deacetylating and inhibiting the NF-κB p65 subunit. In the innate immune system, SIRT2 is the most highly expressed sirtuin in myeloid cells and exerts crucial regulatory effects on macrophage activation and immune homeostasis. | 32768387  25072851 |
| HGF | 0.0792 | 0.0854 | Hepatocyte growth factor promotes differentiation of monocytes into regulatory, IL-10-high/IL-12-low accessory cells that induce FoxP3+ regulatory T cells, and directly suppresses macrophage IL-6 production while enhancing IL-10, indicating a net immunoregulatory/anti-inflammatory function. | 16527888  21072211 |
| IL-33 | 0.0251 | 0.0868 | Nuclear-associated IL-1-family alarmin constitutively expressed in endothelial and epithelial cells; released upon cell injury or necrosis, IL-33 signals through its receptor ST2 (IL-1RL1) to activate mast cells, ILC2s, Th2 cells, and a highly suppressive ST2+ regulatory T cell subset, giving it dual pro-inflammatory (type 2 immunity, tissue remodeling/fibrosis) and tissue-repair/immunoregulatory functions depending on chronicity and cellular context. | 29247993  38082335 |
| MCP-3 | 0.0505 | 0.0993 | CCL7 is a monocyte and eosinophil chemoattractant that synergizes with CXCL8 to enhance neutrophil chemotaxis during inflammation. Plasma MCP-3 rises significantly within 6 to 24 hours post-injury in mice, consistent with a role as an acute-phase reactant that signals tissue damage and initiates immune cell infiltration. | 15075362  34833127 |
