## Supplementary material for "Multiple Organ Dysfunction Trajectory Phenotypes After Severe Trauma Are Associated With Distinct Immune Profiles and Outcomes": SDC4 – Supplemental Figures

**Supplemental Digital Content 4. Supplemental Figures**

**Figure S1: Individual mSOFA trajectories by phenotype.** Cluster phenotypes derived from longitudinal mSOFA scoring. Combined smoothed trajectories with standard error are shown out to 28 days (mSOFA trajectories). Raw data trajectory plots are shown individually across the 5 cluster phenotypes (c1-c5). Cluster sizes: c1 (n=14), c2 (n=22), c3 (n=21), c4 (n=7), c5 (n=5).

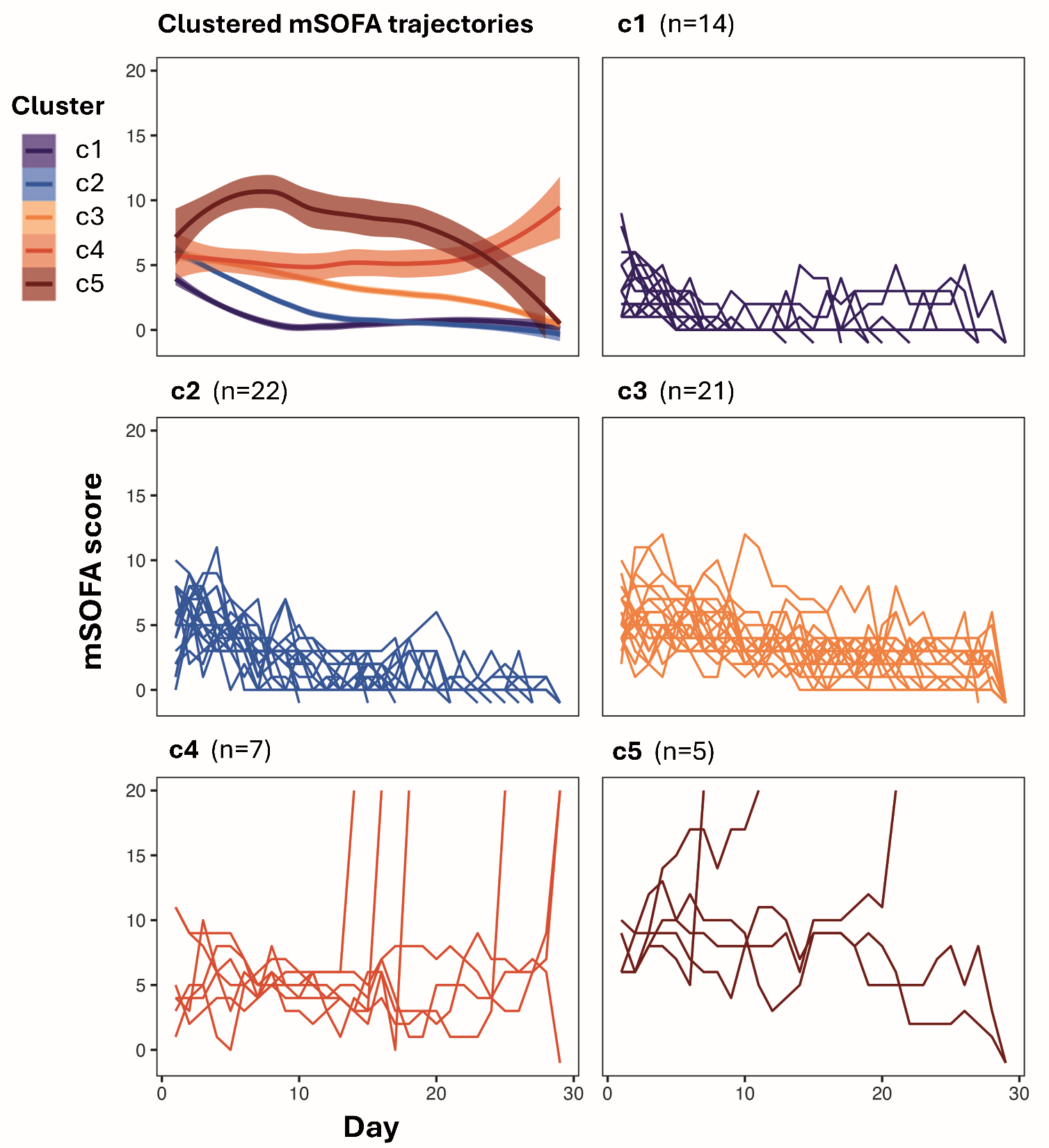

**Figure S2: Comparison of hospital length of stay across lower severity phenotypes (c1/c2/c3).** Hospital length of stay in days was compared across the lower severity phenotypes c1, c2 and c3.

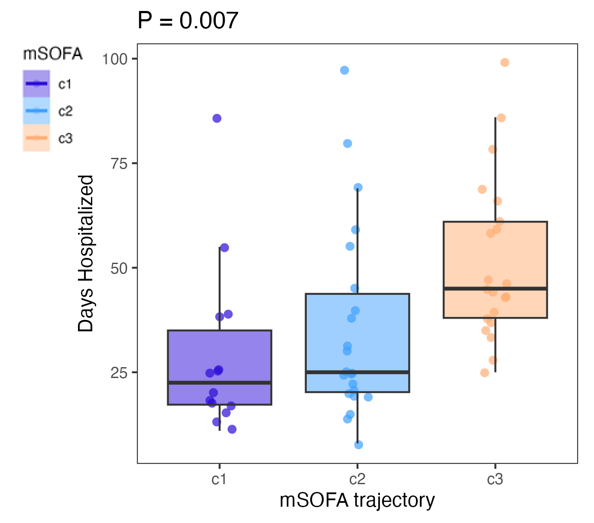

**Figure S3: Longitudinal mSOFA trajectory phenotypes by organ system.** In the top row, smoothed longitudinal organ subscore trajectories with standard error are shown for each mSOFA component organ system (Cardiovascular, coagulation, liver, renal, respiratory) and by cluster phenotype (c1-c5). Subsequent rows show individual organ subscore trajectories by phenotype (c1-c5). Each tracing line is an individual subject. Cluster sizes: c1 (n=14), c2 (n=22), c3 (n=21), c4 (n=7), c5 (n=5).

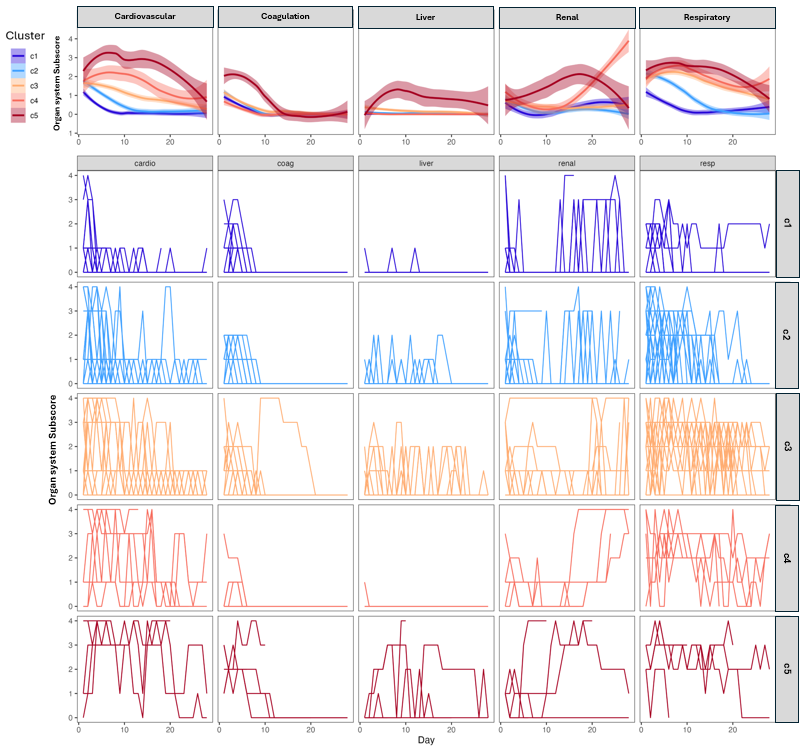

**Figure S4A: Clinical characteristics across phenotypes – Key continuous variables.** Continuous variables of clinical characteristics compared across phenotypes. 1^st^ row: age, Injury Severity Score, initial measured serum lactate, peak lactate through 6 hours, peak lactate through 24 hours. 2^nd^ row: Packed red blood cell volume transfused through 24 hours, whole blood volume transfused through 24 hours, platelets volume transfused through 24 hours, fresh frozen plasma transfused through 24 hours, and crystalloid volume administered through 24 hours. Significant across group and pairwise comparisons (p<0.05) are shown. *, **, ***, are across group and pairwise comparisons with p<0.01. aov, analysis of variance; FDR, Benjamini-Hochberg false discovery rate; P, unadjusted p-value.

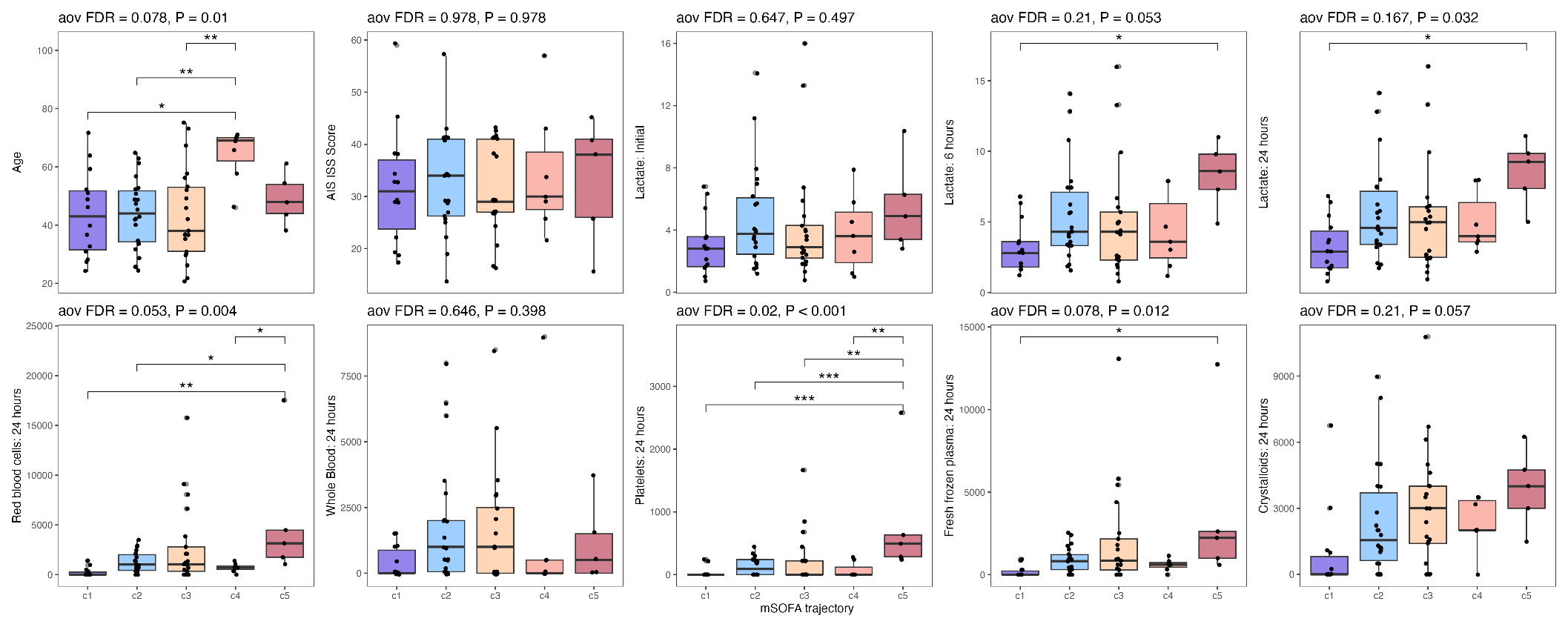

**Figure S4B: Clinical characteristics across phenotypes – Categorical variables.** Categorical variables of clinical characteristics compared across phenotypes. Raw counts for each category are shown within each panel (white square). FDR, Benjamini-Hochberg false discovery rate; P, unadjusted p-value. “inj_crush”, crush injury present; “alcohol_adm”, positive serum ethanol test on admission; “drug_adm”, positive drug screen on admission; “ais_”, ais region ≥3; “pre_transfer”, pre-hospital transfer from another facility within 6 hours of injury.

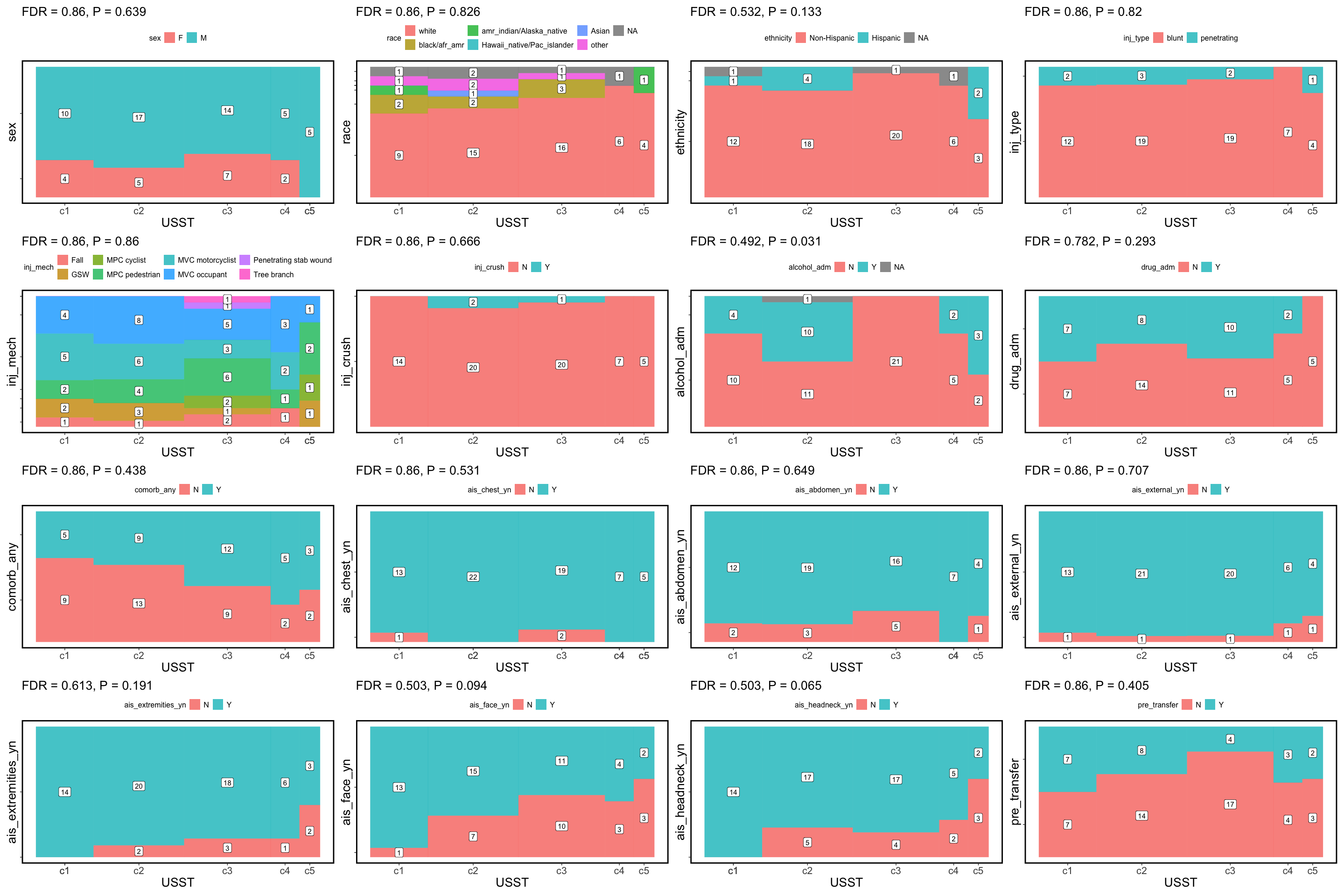

**Figure S4C: Clinical characteristics across phenotypes – All continuous variables.** Continuous variables of clinical characteristics compared across phenotypes as violin plots. aov, analysis of variance; FDR, Benjamini-Hochberg false discovery rate; P, unadjusted p-value. “pre_”, measured pre-hospital; “init_”, 1^st^ hospital measured value; “er_”, measured in emergency department; “_low” ,lowest value measured; “_high”, highest measured value; “hr_”, heart rate; “systolic”, systolic blood pressure; “ais_score”, total injury severity score; “lactate”, serum lactate measurement; “lactate_deficit”, based deficit measurement; “resus_”, resuscitation parameter; “rbc_24”, red blood cell volume transfused - 24 hours; “WB_24”, whole blood transfused - 24 hours; “plat_24”, platelet volume transfused – 24 hours; “xloid_24”, crystalloid administered – 24 hours; “alcohol_adm”, admission blood ethanol level.

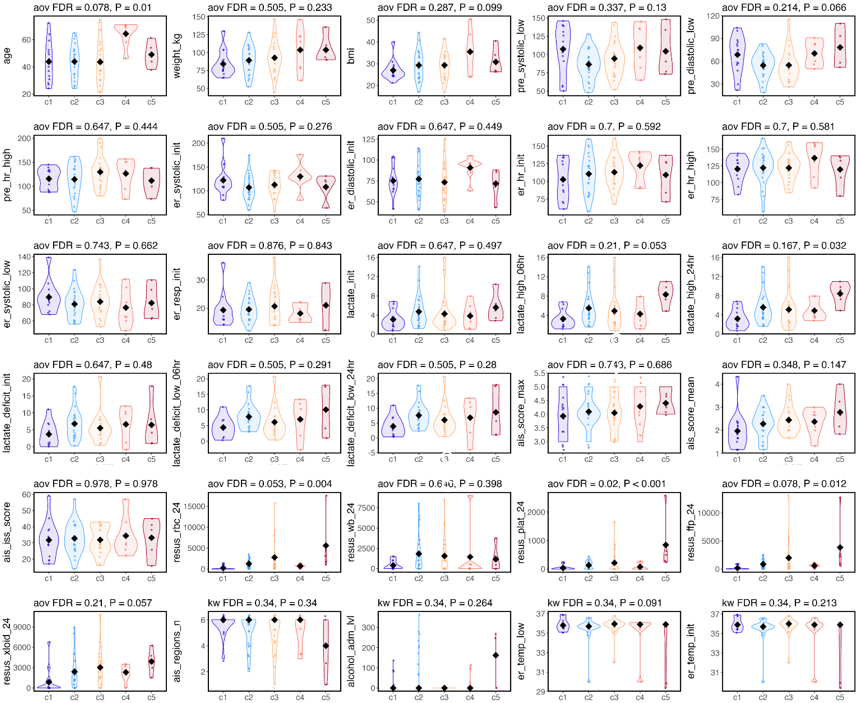

**Figure S5: Top 15 immune marker trajectories that differentiate organ dysfunction phenotypes.** Mixed effects model analysis revealed a total of 40 significant marker trajectories that differed across the 5 mSOFA phenotypes (FDR<0.1). The top 15 immune marker trajectories by FDR are shown in order of significance.

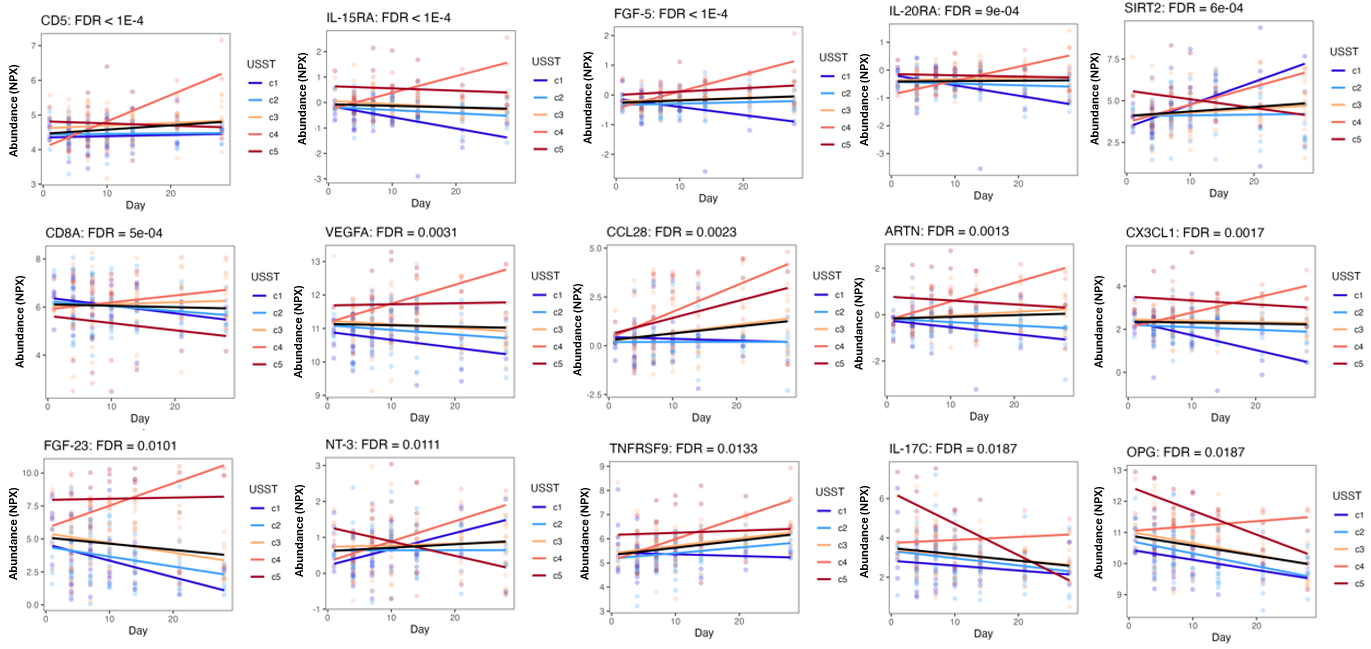

**Figure S6: Distinguishing immune marker trajectories among the lower severity phenotypes (c1/c2/c3).** Select plasma protein immune markers with significant difference in longitudinal trajectory (FDR<0.1) across the c1/c2/c3 phenotypes. The full list of markers meeting FDR<0.1 (n=13) is listed in *SDC3 Table S4*.

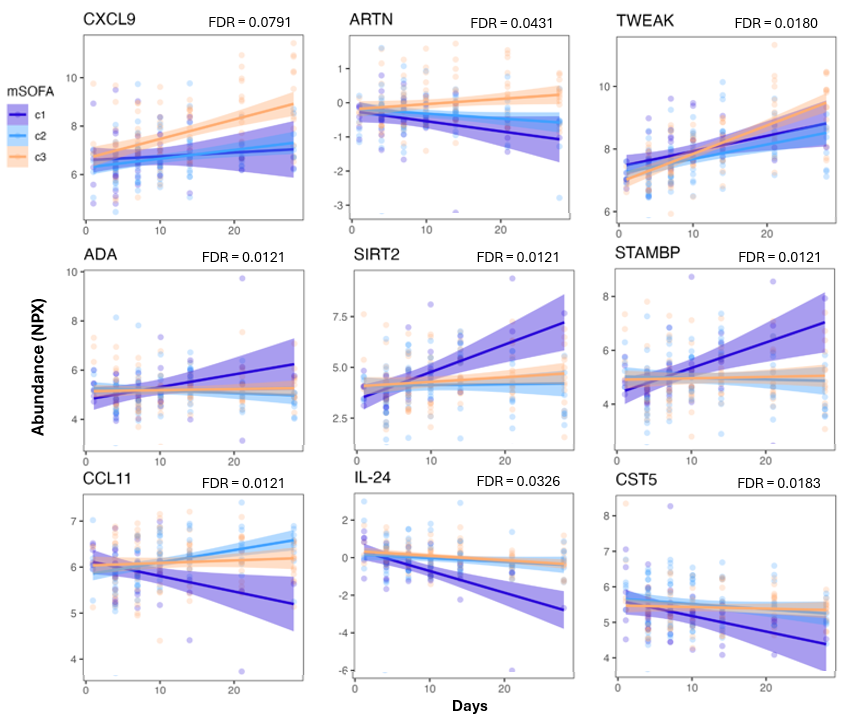
